# Ancestry and somatic profile predict acral melanoma origin and prognosis

**DOI:** 10.1101/2024.09.21.24313911

**Authors:** Patricia Basurto-Lozada, Martha Estefania Vázquez-Cruz, Christian Molina-Aguilar, Amanda Jiang, Dekker C. Deacon, Dennis Cerrato-Izaguirre, Irving Simonin-Wilmer, Fernanda G. Arriaga-González, Kenya L. Contreras-Ramírez, Emiliano Ferro-Rodríguez, Jamie Billington, Eric T. Dawson, J. Rene C. Wong-Ramirez, Johana Itzel Ramos-Galguera, Alethia Álvarez-Cano, Dorian Y. García-Ortega, Omar Isaac García-Salinas, Alfredo Hidalgo-Miranda, Mireya Cisneros-Villanueva, Peter A. Johansson, Héctor Martínez-Said, Pilar Gallego-García, Mark J. Arends, Ingrid Ferreira, Mark Tullett, Rebeca Olvera-León, Louise van der Weyden, Martín del Castillo Velasco Herrera, Rodrigo Roldán-Marín, Helena Vidaurri de la Cruz, Luis Alberto Tavares-de-la-Paz, Diego Hinojosa-Ugarte, Rachel L. Belote, D. Timothy Bishop, Marcos Díaz-Gay, Ludmil B. Alexandrov, Yesennia Sánchez-Pérez, Gino K. In, Richard M. White, Patrícia A. Possik, Robert L. Judson-Torres, David J. Adams, Carla Daniela Robles-Espinoza

## Abstract

Acral melanoma, which is not ultraviolet (UV)-associated, is the most common type of melanoma in several low- and middle-income countries including Mexico. Latin American samples are significantly underrepresented in global cancer genomics studies, which directly affects patients in these regions as it is known that cancer risk and incidence may be influenced by ancestry and environmental exposures. To address this, we characterise the genome and transcriptome of 123 acral melanoma tumours from 92 Mexican patients, a population notable because of its genetic admixture. Compared with other studies of melanoma, we found fewer frequent mutations in classical driver genes such as *BRAF*, *NRAS* or *NF1*. While most patients had predominantly Amerindian genetic ancestry, those with higher European ancestry had increased frequency of *BRAF* mutations and a lower median number of structural variants. The tumours with activating *BRAF* mutations have a transcriptional profile more similar to cutaneous non-volar melanocytes, suggesting that acral melanomas in these patients may arise from a distinct cell of origin compared to other tumours arising in these locations. *KIT* mutations were found in a subset of these tumours, and quadruple wild-type samples (non *BRAF/NRAS/NF1/KIT*) differed from mutated samples in their structural genomic profile and overall and recurrence-free survival patterns. Transcriptional profiling defined three expression clusters; these characteristics were associated with recurrence-free and overall survival. We highlight potential novel low-frequency drivers, such as *PTPRJ*, *NF2* and *RDH5*. Our study enhances knowledge of this understudied disease and underscores the importance of including samples from diverse ancestries in cancer genomics studies.

## Introduction

Melanoma is classified into several clinicopathological subtypes based on tumour site of presentation and histopathological features. Acral melanoma (AM) is an understudied melanoma subtype due to its low incidence globally, and because it represents a small proportion of melanoma cases in European-descent populations^1,2^; however, AM represents the vast majority of melanoma cases in some Latin American, African and Asian countries due to the lower incidences of ultraviolet (UV)-induced melanoma subtypes^3^. Additionally, the causes of this type of disease are unknown, with patients managed in a similar way to UV-associated cutaneous melanoma (CM). However, its site of presentation and genomic characteristics are vastly different^4^.

AM arises on the glabrous (non-haired) skin of soles, palms and in the nail unit (subungual location), and its genome differs significantly from other CM subtypes^5^. In contrast to UV-linked subtypes like superficial spreading or lentigo maligna melanoma, AM has a lower burden of single nucleotide variants (SNVs), a higher burden of structural variants, and a low prevalence of mutational signatures SBS7a/b/c/d, which are associated with UV irradiation^6–10^. Genes that are frequently mutated in CM such as *BRAF*, the *RAS* genes and *NF1*, are reported to be altered at a significantly lower frequency in AM. This, coupled with the comparatively lower number of studies of AM when compared to other CM subtypes, has translated into limited available therapies for AM management.

It is known that cancer risk and incidence, as well as tumour genomic profiles, vary with ancestry and geographical location^11–13^. Since most genomic studies on AM have been performed on patients of European or Asian ancestry, we considered it necessary to examine the genomics of this subtype of melanoma in Latin Americans. Specifically, Latin American populations have been substantially underrepresented in cancer genomic studies, with only about 1% of all samples in cohorts such as the Pan-Cancer Analysis of Whole Genomes (PCAWG), the Cancer Genome Atlas (TCGA) and other repositories, and those contributing to cancer genome-wide association studies (GWAS), being of Latin American origin^14–16^. Identification of differences in the genomic profile among populations can potentially aid the discovery of germline/inherited or environmental factors related to AM aetiology, as well as identify optimal therapeutic strategies for all patients.

In this study, we analysed 123 AM samples from 92 Mexican patients through genotyping, exome sequencing, SNV and insertion/deletion (indel) variant calling, copy number estimation, and gene expression profiling, and examined the correlation of these molecular characteristics with clinical variables. We reveal a significant correlation between genetic ancestry and *BRAF* somatic mutations, as well as a distinct transcriptomic profile in these tumours compared to non-*BRAF* mutated samples. We also identify significant differences in recurrence-free and overall survival among patients with driver mutations compared to patients with no mutations in driver genes, as well as among patients with tumours with distinct gene expression profiles.

## Results

### Ancestry and clinical characteristics of Mexican AM patients

A total of 123 uniformly ascertained samples from 92 patients from a large Mexican tertiary referral hospital were analysed in this study (**Methods**, **Supplementary Table 1**). Eighty-nine of these tumours were primaries, 27 were metastases, five were recurrences, one was an in transit lesion, and one was unknown (**Supplementary Table 1**). Latin American genomes are generally a mixture of European, African and Amerindian ancestry. Of note, 90% of genotyped samples (n=80) in this study had predominantly Amerindian ancestry (median 81%) (**Supplementary Figure 1, Supplementary Table 2**) with European and African ancestries contributing a median of 13.6% and 2.5%, respectively. The median age of the patients in this cohort was 60, with 59% of the patients being female. Most patients were stage III (AJCC 8th edition)^17^ at diagnosis, and the most common primary site was the foot, most frequently the sole. The median Breslow thickness was 4.0mm and most primary tumours were ulcerated (68%) (**Table 1**). It should be noted that only four patients received immune check point inhibitors or targeted therapy, due to lack of access.

**Table 1.** Clinical information for patients included in this study.

|  |  |  |
| --- | --- | --- |
| **Number of patients** |  | 92 |
| **Age** |  |  |
|  | *Median* | 60 |
|  | *Mean* | 61.22 |
|  | *Min* | 32 |
|  | *Max* | 98 |
| **Sex** |  |  |
|  | *Female* | 54 |
|  | *Male* | 38 |
| **Stage** |  |  |
|  | *IV* | 9 |
|  | *III* | 47 |
|  | *II* | 25 |
|  | *I* | 10 |
|  | *In situ* | 1 |
| **Ulceration** |  |  |
|  | *Yes* | 61 |
|  | *No* | 29 |
|  | *NA* | 2 |
| **Site** |  |  |
|  | *Feet* | 76 |
|  | *Hands* | 4 |
|  | *Subungual* | 12 |
| **Breslow Thickness** |  |  |
|  | *Median* | 4 |
|  | *Mean* | 6.61 |
|  | *Min* | 0.45 |
|  | *Max* | 50 |

### Genomic profiling of AM samples identifies correlations of sex and ancestry with somatic alterations

Considering all 123 samples, AM tumours showed a SNV+indel [hereinafter referred to as tumour mutational burden (TMB)] mean of 0.95 mutations per megabase (mut/Mb) and a median of 0.87 mut/Mb (range: 0-3.49 mut/Mb). When including only one sample per patient, with primaries being preferentially selected, the most frequently mutated genes were *NRAS* (14% of samples, *q*-value < 4.97×10^−10^), *KIT* (14% of samples, *q*-value = 4.97×10^−10^), *BRAF* (13%, *q*-value=3.86×10^−7^) and *NF1* (9%, *q*-value=0.0001) (**Figure 1a, Supplementary Table 3**). For *BRAF*, all mutations except one were V600E, with one L597R (**Figure 1b**). These genes were identified as being under positive selection (**Methods**) and represent known driver genes. These genes showed the characteristic mutational profile of oncogenes with a predominance of hotspot missense mutations, except for *NF1*, which showed a pattern characteristic of a tumour suppressor and had large deletions (an exon or larger), frameshift insertions, deletions, splice donor and nonsense mutations distributed throughout (**Figure 1b**). Notably, these genes exhibit mutual exclusivity (only one patient has tumours with mutations in more than one of these genes) which likely reflects their functional redundancy in activating the MAPK pathway. Separate capillary sequencing of the *TERT* promoter in 76 samples belonging to 64 patients identified that six carry the -124 promoter mutation (9.3%) and two out of 59 patients for which the -146 position was successfully amplified carry a mutation in this position (3.4%) (**Supplementary Table 4**). In total, we estimate that 10.5% of patients have an activating *TERT* promoter mutation, which is similar to estimates in other studies^7,18^. All samples from all patients that had multiple samples sequenced and that could be assessed had a concordant *TERT* genotype, in agreement with an early emergence of this mutation during tumour evolution^18^. Other genes previously reported as mutated in other melanoma subtypes, as well as other cancer types are also mutated in this cohort, such as *TP53*, *HRAS* and *KRAS* (**Figure 1a, Supplementary Figure 2**). In summary, the “classic” melanoma driver genes (*N/H/KRAS*, *BRAF* and *NF1*) are mutated in 40% of Mexican AM samples, with most of the samples in this cohort therefore being classified as “triple wild type” melanomas. Apart from the known *HRAS*, *SPRED1*, *TP53*, and *KRAS* driver genes, we also find mutations in *PTPRJ*, *ATM*, *NF2* and *RDH5* (**Supplementary Figure 2a,b**). Specifically, in those tumours without mutations in any of the abovementioned four driver genes [*BRAF*, *NRAS*, *KIT*, *NF1*, “quadruple wild-type” (QWT)], we find two tumours each from different patients with deleterious mutations in *ATM* and *RDH5* (**Supplementary Figure 2b**). The mutations in these genes are all protein-changing and deleterious. All these genes have previously been linked to tumour suppressor activities in either acral or mucosal melanomas^19–24^, as well as other cancer types, and may represent low-frequency drivers. We also observe a significantly higher proportion of women carrying mutations in driver genes vs men (Two-tailed Fisher’s test *P*=0.003) (**Supplementary Table 5**). After adjusting for date of diagnosis, age at diagnosis, ancestry and tumour stage, the Odds Ratio (OR) of having a mutation in a driver gene in female patients (compared to men) was estimated to be 3.83 (95% CI 1.32, 11.07) (multivariate logistic regression, *P* = 0.013).

**Figure 1.**
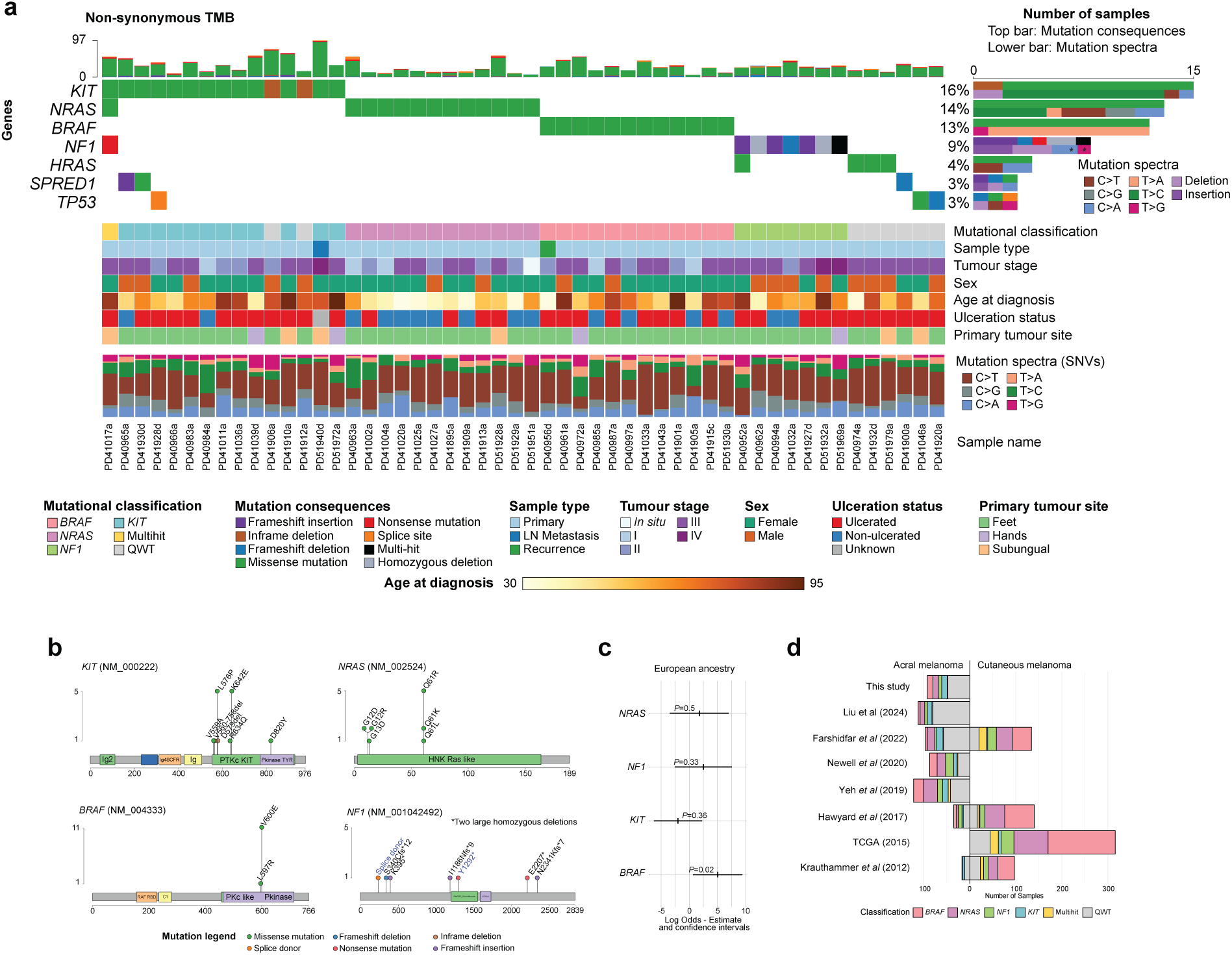
Somatic landscape of acral melanoma in Mexican patients. A) Oncoplot depicting the 7 most mutated genes according to dNdScv and their status in the samples with mutations in these (52 samples out of 92, one per patient). Mutational classification, sample type, tumour stage, sex, age at diagnosis, ulceration status, tumour site and mutational spectra are shown by sample. B) Mutations found in *KIT*, *NRAS*, *BRAF* and *NF1*, which are the most significantly mutated genes. C) A logistic regression model controlling for age, sex, and TMB was fitted to predict the presence or absence of a mutation on the AM samples using the inferred ADMIXTURE cluster related to the European ancestry component. Log odds estimate and confidence intervals are depicted for the four driver genes. D) Barplot depicting the number and mutational classification of samples in different acral and cutaneous melanoma studies.

When examining the relationship between ancestry and somatic profile, we identified significantly higher odds (*P*-value=0.02) of carrying a *BRAF* somatic mutation with increasing European ancestry in a linear model controlling for age at diagnosis, sex, and TMB (**Figure 1c**). Patients with mutations in *KIT* showed a tendency for higher Amerindian ancestry (**Figure 1c**, **Supplementary Figure 3**). We also found that patients with *NRAS* mutations were younger at diagnosis (median and mean age of diagnosis for patients with *NRAS* mutations = 49 and 50.84 vs without = 63 and 62.9, respectively), but this effect is likely mediated by ulceration status, as patients with *NRAS* mutations have a significantly lower rate of ulceration (Two-tailed Fisher’s Exact test *P*=0.016).

Out of 22 patients for whom we sequenced at least two different samples (*e.g*., a primary and a metastasis), 13 have mutations in *NRAS/KIT/BRAF/NF1*. For *BRAF*, all four patients have mutations across all samples, and for *NRAS*, three out of four patients have a *NRAS* mutation in both the primary and lymph node metastasis. These data suggest that these mutations appear as an early event in tumour evolution. For patients with *KIT* mutations, about half of the times the mutation is found in the primary and lost in the metastasis. These data agree with Wang *et al* (2024)^18^, and suggest that metastases are seeded before the appearance of these mutations (**Supplementary Table 6**).

Collectively, these results are similar to those reported in other acral melanoma studies^6,7,10,25–28^, with some important differences: First, the genetic composition of the patients in our study includes a significant proportion of Native American ancestry, which is severely underrepresented in already published cancer genomics studies and permits the identification of relationships of specific ancestries with somatic characteristics. In addition, the fraction of activating *BRAF* mutations is lower than in the studies with predominantly European-descent patients, and more similar to those with Chinese patients, probably due to the positive relationship between *BRAF* mutation and European ancestry (**Figure 1d**).

### Somatic copy number landscape of AM samples identifies correlations with somatic alterations

Somatic copy number alteration (SCNA) analysis across all samples showed a higher burden of amplifications than deletions (**Figure 2a**). Examination of 47 samples, one per patient, that passed our stringent quality filtering for this type of analysis (**Methods**), showed that 18 regions were significantly amplified, and six regions were frequently deleted. About a quarter (11, 23%) of these 47 samples had whole genome duplication (WGD) events. Potential driver genes in frequently amplified regions include *TERT* (43%), *CRKL* (36% of samples), *GAB2* (30%) and *CCND1* (28%) (**Supplementary Tables 4,7-9**). Regions that showed recurrent deletions contained genes such as *CDKN2A*, *CDKN2B, ATM* and *TP53*. Specifically, *CDKN2A* and *CDKN2B* had deletions in 57% of samples, with only two of them (out of 27) being homozygous deletions, while *ATM* and *TP53* both had deletions in 38% and 34% of samples, respectively. No association was found between ancestry and any of the significantly altered CNA regions.

**Figure 2.**
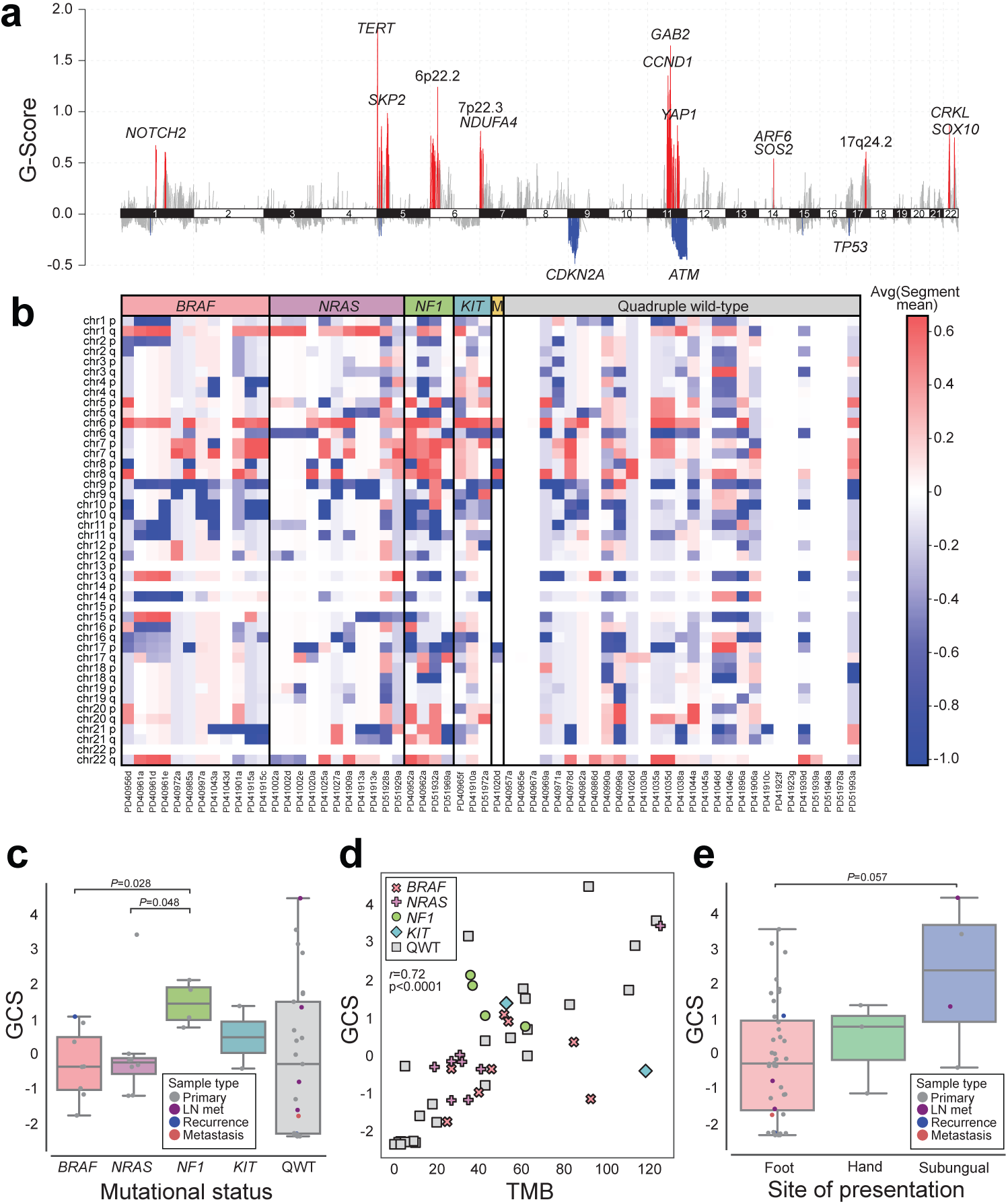
DNA copy number landscape of acral melanoma and molecular and clinical correlates in Mexican patients. A) Regions of amplification (red) and deletion (blue) in 47 acral melanoma samples, one per patient, as identified by GISTIC2. Known drivers, or the chromosomal regions, are shown. This analysis shows alteration with respect to the normal sample, *i.e*., with respect to a ploidy of 2. B) Heatmap showing regions of amplification (red) and deletion (blue) by sample and chromosomal arm in all 60 samples classified into genomic subgroups. This analysis shows alterations with respect to the estimated tumour sample ploidy. C) Boxplot of global copy-number scores (GCS) of 47 samples, one per patient, classified by genomic subgroup. *P*-values are from Wilcoxon-Mann-Whitney paired tests. D) Scatter plot of TMB (X axis) and GCS (Y axis) for 47 samples, one per patient. Dots represent samples, coloured by genomic subtype. Pearson’s product-moment correlation coefficient and associated *P*-value is shown. E) Boxplot of GCS of 47 samples, one per patient, classified by tumour site. GCS scores are calculated with respect to tumour sample ploidy. For box plots, the central line within each box represents the median value, the box boundaries represent the interquartile range (IQR), and the whiskers extend to the lowest or highest data point still within 1.5xIQR.

SCNA profiles varied depending on whether samples had mutations in driver genes or were QWT. Specifically, samples with mutations in driver genes (n=22) showed preferential amplification of *NOTCH2* (*P*=0.021, one-sided exact Fisher test), whereas *CCND1* (*P*=0.044), *ARF6* and *SOS2* (both in same amplification peak, *P*=0.043, all one-sided exact Fisher tests) were preferentially amplified in QWT tumours (n=25). The 8p12 region, containing genes such as *FGFR1* and *TACC1*, was only found amplified in five QWT samples, whereas regions 11q13.4 and 6p24 were found amplified along several deleted regions in mutated tumours (**Supplementary Tables 10-15, Supplementary Figures 4-5**).

When stratifying samples by mutational status (considering *BRAF*-, *NRAS*-, *NF1*-, *KIT*-mutated and multi-hit, which included one sample with mutations in more than one of these drivers), we saw statistically significant differences in SCNA among groups, specifically, *NRAS*- and *BRAF*-mutated tumours had significantly fewer SCNAs (Global copy number alteration score [GCS], **Methods**) than *NF1*-mutated tumours (**Figure 2b-c**). Samples without mutations in these drivers had a range of GCS scores. Considering all samples, those with *NRAS* and *BRAF* mutations had the lowest median TMB, while *KIT*-mutated and multi-hit tumours had the highest median TMB (**Supplementary Figure 6**). We found a significant correlation between GCS score and TMB (Pearson’s product moment correlation coefficient=0.72, *P*-value<0.0001) (**Figure 2d**). Tumours from the subungual region also had a higher median GCS score than those found on the feet (**Figure 2e**).

### Mutational signature analyses identify potential sources of mutation

Single-base substitution mutational signature analysis across 116 samples that carried at least one SNV revealed previously reported COSMICv3.4 signatures SBS1, SBS5, and SBS40a (**Supplementary Figure 7**). Apart from clock-like signatures SBS1 and SBS5^29^, SBS40a was also prevalent across the cohort, contributing 53.38% of mutations to the total (**Supplementary Figure 8**). SBS40a is of unknown origin but was originally identified in kidney cancer and is present in many cancer types^30^. Copy number signature analysis identified a number of previously reported signatures across different samples (**Methods**, n=60 samples)^31,32^. CN1, which has been associated with a diploid state and CN9, which is potentially caused by local loss of heterozygosity on a diploid background, dominated the CN landscape (**Methods, Supplementary Figure 7**). As expected, signatures related to chromothripsis (CN7, CN8) were also found in several samples across the cohort. The number of indels in the samples was too low to add meaningful information (average: 2.52 indels per sample), so signature analysis for indels was not performed. We similarly did not find any significant associations between ancestry and any mutational signature. Nevertheless, this analysis is limited by the small numbers of mutations and the formalin-fixed paraffin-embedded (FFPE) origin of these samples.

### Acral melanomas with activating BRAF mutations exhibit a transcriptional signature more characteristic of non-acral cutaneous melanomas

It has been previously postulated that *BRAF*-mutated acral melanomas might be more biologically similar to melanomas from non-acral sites than to other acral melanomas^10,33^, and, due to the observed correlation of European ancestry with *BRAF* mutation rate, we decided to investigate this hypothesis. We successfully extracted and sequenced RNA from 77 primary tumours from different patients (**Supplementary Table 1, Methods**). We then generated a gene signature-based score for identifying acral-versus cutaneous-derived melanomas. For this, we sourced a list of candidate genes from AM and CM datasets (**Methods, Supplementary Table 16**) and identified twenty genes with high classification accuracy in a training cohort of 10 primary AMs (used to derive a v-mel score, or “A” for acral) and 10 primary CMs (used to derive a c-mel score, or “C” for cutaneous) recruited at the University of Utah (**Figure 3a-b**). We then obtained scores (v-mel/c-mel, or A:C) for samples in our dataset of AMs, separating primary *BRAF*-activating (n=10) vs *BRAF*-wildtype (n=67) tumours. We observed a difference between *BRAF*-activating and *BRAF*-wildtype tumours (*P*-value=0.045), with *BRAF*-activating tumours having a score closer to CMs (**Figure 3c**). We then replicated this analysis in an independent cohort of 63 AMs from Newell *et al* (2020)^7^ (*BRAF*-activating n=10, wild-type n=53), which further confirmed these results (*P*-value=0.039) (**Figure 3d**). Therefore, we explored the possibility that this difference could be due to downstream mutated *BRAF* signalling. First, we replicated this analysis in the TCGA cutaneous melanoma data, finding no significant differences among *BRAF* and non-*BRAF* mutated samples (**Supplementary Figure 9a**). We then examined datasets in which mutant *BRAF* was introduced into primary melanocytes in a doxycycline-inducible manner^34^. We found that the c-mel signature genes were not activated downstream of mutant *BRAF*, further suggesting that the classifier does not simply reflect *BRAF*-driven transcriptional changes (**Figure 3d**). Using a recently developed method for assessing gene signature similarity^35^, we compared the c-mel gene signature from our classifier to a previously published set of genes directly activated by mutant *BRAF* in melanoma cells^36^. We found no significant correlation between these signatures (**Supplementary Figure 9b**).

**Figure 3.**
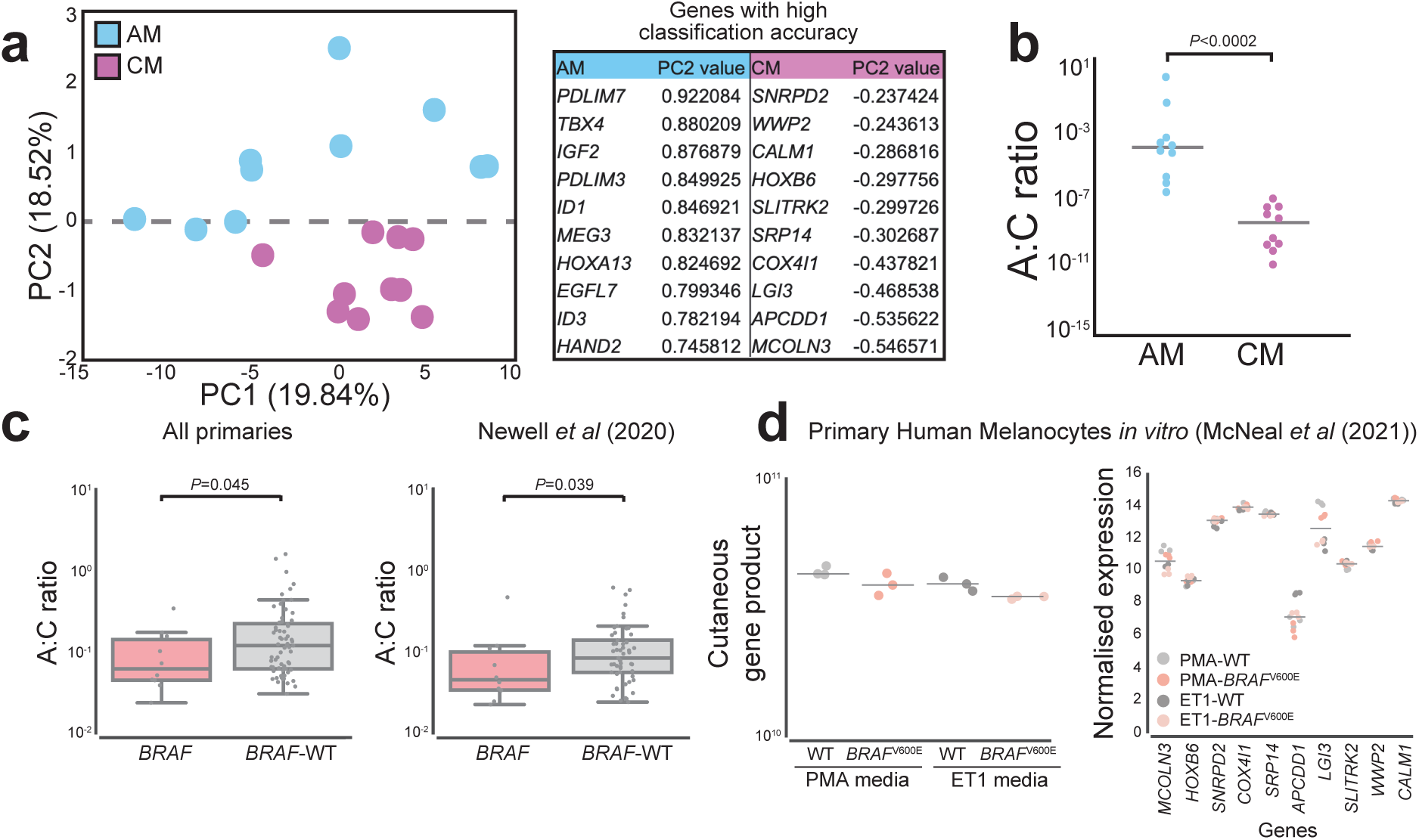
Comparisons of the transcriptional profile of *BRAF*-activating and *BRAF*-wildtype AM tumours. A) Elucidation of genes used to classify acral vs cutaneous melanoma samples. PCA of acral melanoma (blue) and cutaneous melanoma (purple) samples (left panel). Loadings on PC2 were used to identify the top differentially expressed genes contributing to the variance between acral melanomas and cutaneous melanomas (right panel). B) Scatter plot showing the distribution of the acral:cutaneous (A:C) gene expression ratios between test acral and cutaneous melanoma samples. AM samples are represented by blue dots, and CM samples are represented by purple dots (*P*-value < 0.0002). c) Left panel. Comparison of A:C gene expression ratio in AM samples with different mutation status. Box and whiskers plot comparing two groups: *BRAF*-WT and *BRAF*-activating mutated tumours. Right panel. Comparison of A:C gene expression ratio in AM samples with *BRAF-*activating mutations and *BRAF*-WT tumours from Newell *et al* (2020)^7^. The central line within each box represents the median value, the box boundaries represent the interquartile range (IQR), and the whiskers extend to the lowest or highest data point still within 1.5xIQR. Individual data points are plotted as dots. Statistical significance was assessed using individual Wilcoxon Mann-Whitney tests. D) Left panel. Comparison of the product of the cutaneous genes in normal human melanocytes. Melanocytes were cultured in PMA or ETA containing media with or without doxycycline-induced *BRAF*^V600E^ expression. Each dot is an individual biological replicate with horizontal lines indicating median values. Right panel. Relative expression levels of cutaneous genes across individual normal human melanocytes. Melanocytes were cultured in PMA or ETA containing media with or without doxycycline-induced *BRAF*^V600E^ expression. Each point represents a biological replicate with horizontal lines indicating median values.

In conclusion, in these comparisons, *BRAF*-activating tumours expressed a more “CM-like” transcriptional program, indicating that *BRAF*-mutated melanomas that occur at acral sites are transcriptionally closer to non-acral cutaneous melanomas, a transcriptional program that is not explained by *BRAF* downstream signalling and are associated with increasing European genetic ancestry.

### Transcriptional landscape of AM tumours identifies three subgroups with distinct clinical and prognostic characteristics

We then applied a more stringent quality filter, including coverage and alignment features, to primary tumours in this collection with 44 samples remaining for further analyses (**Methods**, **Supplementary Table 1**). Consensus clustering of gene expression identified three sample groups with distinct transcriptional profiles (**Methods**, **Figure 4a-b, Supplementary Table 17**). Cluster 1 was characterised by an epidermal/immune profile, with high expression of keratin genes (*e.g*. *KRT1*, *KRT9*), cytokines (*e.g*., *CXCL16*, *IL7, IL4R, IL1R, IL15RA, CXCL14*), and processes such as epidermis development, cell-cell junction organisation and wound healing (**Supplementary Figure 10, Supplementary Table 18**); Cluster 2 was characterised by a mitotic/proliferative-related signature, with high expression of genes such as *MITF* and *TYR,* and processes such as chromosome segregation, nuclear division and mitochondrial translation (**Supplementary Figure 11, Supplementary Table 19**), and Cluster 3 showed expression mostly of respiration and oxidative phosphorylation-related genes and some pigmentation-related genes (e.g. *SOX10*, *DCT*) (**Supplementary Figure 12, Supplementary Table 20**). Interestingly, Cluster 1 was associated with better prognostic clinical characteristics, such as a smaller proportion of ulcerated samples (53%, vs 93% in Cluster 2 and 57% in Cluster 3), a tendency for earlier clinical stages, and lower mitotic rates (Mann-Whitney U C1 vs C2 *P*-value: 0.041, C1 vs C3 *P*-value: 0.052) (**Figure 4c**). Deconvolution of gene expression profiles also indicated differences in immune cell infiltration composition, with Cluster 1 having a higher proportion of CD4+ T cells and cancer-associated fibroblasts (CAFs), and Cluster 2 having a higher proportion of B-cell infiltration (**Figure 4d-f, Supplementary Figure 13, Supplementary Table 21**).

**Figure 4.**
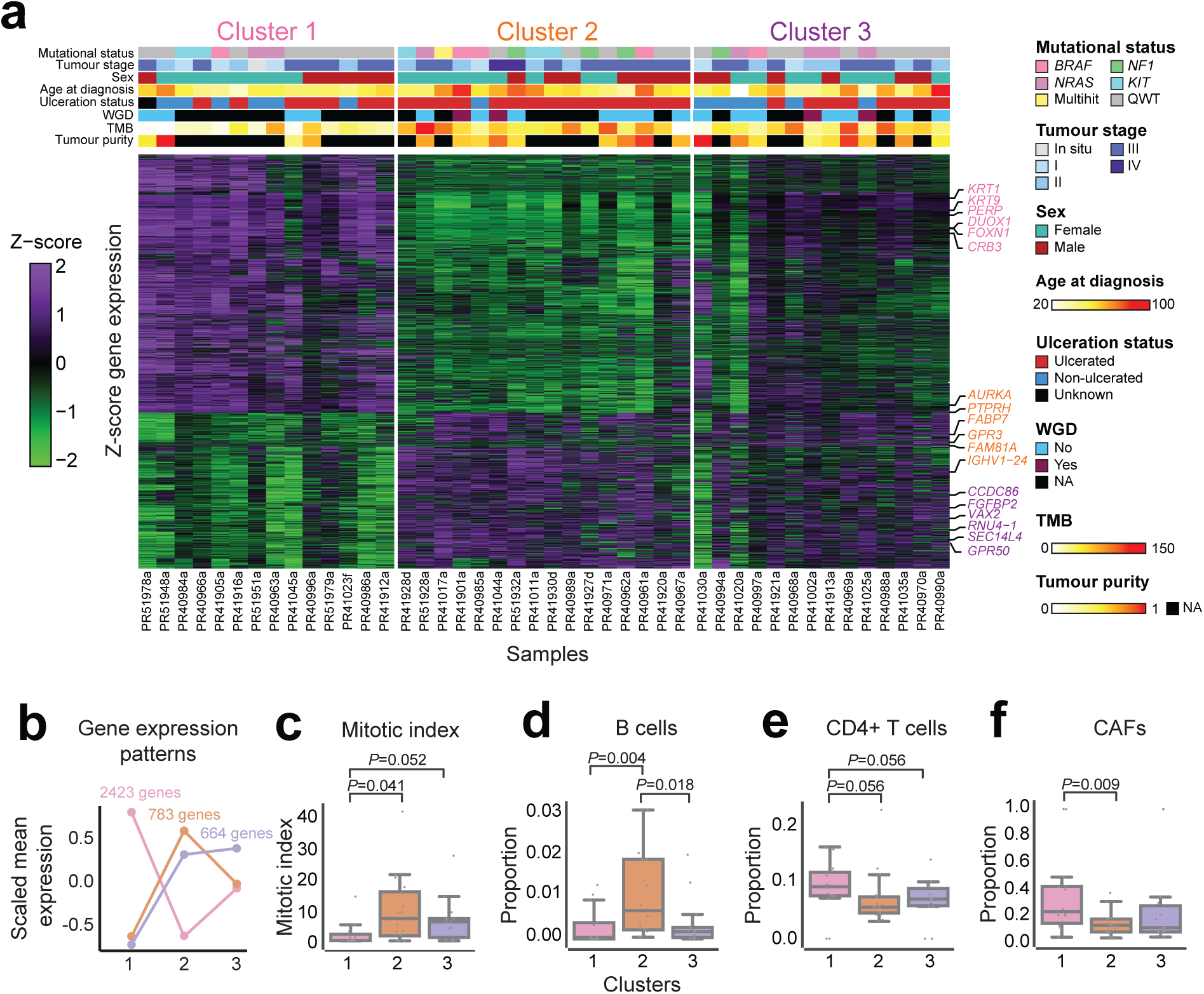
Unsupervised gene expression clustering of primary acral melanoma samples from Mexican patients identifies three main groups. A) Gene expression heatmap showing the 3,870 genes identified as differentially expressed among sample clusters. Samples are in the X axis and genes are in the Y axis. Mutational status and clinical covariates by sample are shown above the heatmap. B) Scaled mean expression patterns per cluster for the three ‘gene programs’ defining each cluster. C) Box plot of mitotic index (Y axis) per sample classified by transcriptional cluster. C) Box plot of B cell proportion (Y axis), as calculated by deconvolution, per sample classified by transcriptional cluster. D) Box plot of CD4+ T cell proportion (Y axis), as calculated by deconvolution, per sample classified by transcriptional cluster. E) Box plot of cancer-associated fibroblasts (CAFs, Y axis), as calculated by deconvolution, per sample classified by transcriptional cluster. The central line within each box represents the median value, the box boundaries represent the interquartile range (IQR), and the whiskers extend to the lowest or highest data point still within 1.5xIQR. Individual data points are plotted as dots. Wilcoxon-Mann-Whitney paired tests were performed.

### Somatic and gene expression profile influence overall and recurrence-free survival

Next, we evaluated whether the genomic and transcriptomic characteristics had any impact on patient overall or recurrence-free survival. We included in the analysis those participants whose primary could be analysed (n=85, **Methods**). The mean time between diagnosis and recruitment was 2.06 years, including 21 participants recruited within 6 months; the range was from a few days to over 10 years. Among these participants, twelve primary tumours had an *NRAS* mutation, eleven had a mutation in *KIT*, eleven had a *BRAF* mutation, seven had *NF1* mutations, one had multiple hits and 43 were classified as wild-type.

Carrying any driver mutation was not associated with age at diagnosis or tumour stage (data not shown). Having a tumour with a driver mutation was, however, associated with a reported recurrence, with 66.7% of mutated tumours having a recurrence as compared to 37.2% of QWT tumours (Pearson two-tailed Chi-squared test *P* =0.007). After adjusting for date of diagnosis, sex, age at diagnosis, ancestry and tumour stage (n=73, primaries with ancestry information available), the OR for a mutated tumour having a recurrence compared to QWT tumours was OR = 5.31 (95% CI 1.56, 18.12), (multivariate logistic regression, *P* = 0.008) (**Figure 5a**). Notably, among the mutated tumours, for each different gene, tumour recurrence was increased over QWT tumours (**Figure 5a**, **Supplementary Table 22**), most notably for *NF1* where all seven of the mutated tumours recurred.

**Figure 5.**
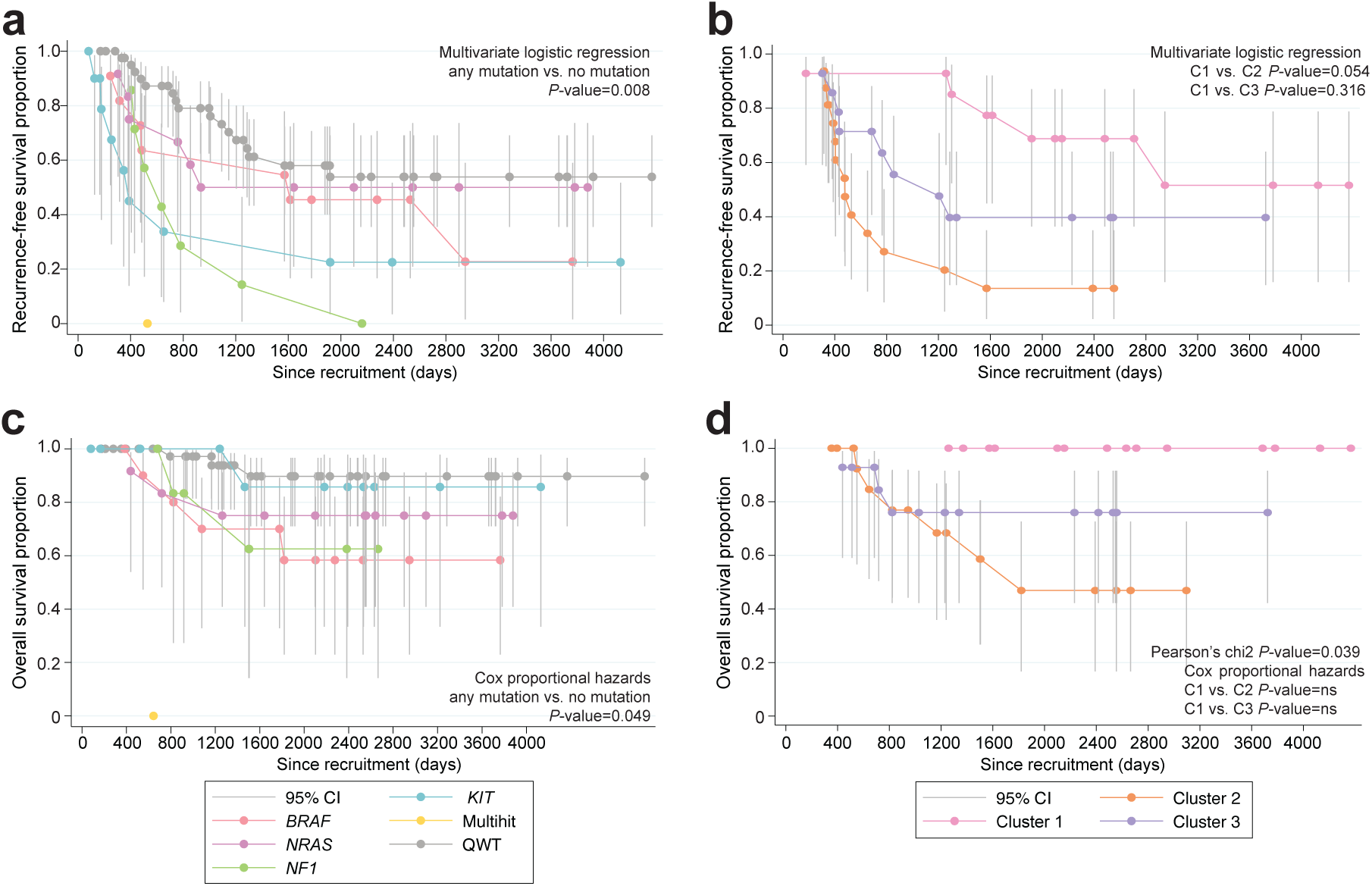
Kaplan-Meier plots of overall and recurrence-free survival for patients by tumour mutational and transcriptional status. A) Recurrence-free survival of patients with and without driver mutations, depicted by each category of the mutational classification. B) Recurrence-free survival for patients with tumours in each of the three transcriptional clusters. C) Overall survival of patients with and without driver mutations, depicted by each category of the mutational classification. D) Overall survival for patients with tumours in each of the three transcriptional clusters. Each panel indicates the test that the depicted *P*-values comes from.

Overall, 44 of the tumours could be classified transcriptomically into one of the three clusters. There was no association between tumour driver mutation and transcriptomic cluster (data not shown). There was, however, evidence of differences in recurrence frequency by cluster, with 35.7% of cluster 1 tumours, 81.2% of cluster 2 tumours and 57.1% of cluster 3 tumours having a recurrence (Fisher’s Exact Test, *P* = 0.04 for homogeneity). Logistic regression adjusting for age at diagnosis, sex, diagnosis date and stage at presentation showed that those tumours from cluster 2 had a higher rate of recurrence as compared to cluster 1 (OR = 6.68, 95%CI 0.97, 46.3), multivariate logistic regression, *P* = 0.054, **Supplementary Table 23**) while cluster 3 had intermediate rates of recurrence (**Figure 5b**).

Fourteen participants (16.5%) died during the study period. Seven percent of participants with QWT tumours and 26.2% of participants with tumours with driver mutations died (*P* = 0.017 for homogeneity). Log rank analysis of time to death from diagnosis showed increased risk of death among those with any mutation (n=73, primaries with ancestry information available) (*P* = 0.036) (**Supplementary Table 24**) while similar analysis by specific mutation showed more extreme significance (*P* < 0.0001) (**Supplementary Table 25**). Cox proportional hazards analysis adjusting for age, sex, tumour stage and ancestry indicated participants whose tumour had any mutation in a driver gene had a significant increase in death in keeping with the analysis of recurrence (HR =5.07, 95% CI 1.02, 25.17, *P* = 0.047) (**Supplementary Table 26**) (**Figure 5c)**.

Finally, survival analysis based on the 44 tumours with transcriptomic classification showed significant variation between the clusters with 0% of cluster 1, 37.5% of cluster 2 and 21.4% of cluster 3 having died (Pearson two-tailed Chi-squared *P*=0.04), again in keeping with the analysis of recurrence, a known major risk factor for survival (**Supplementary Table 27**, **Figure 5d**). Cox proportional hazards analysis adjusting for age at diagnosis, sex, stage at presentation and ancestry did not provide significant evidence of differences between the clusters in terms of mortality rates in keeping with the limited sample size.

## Discussion

In this study, we report the analysis of the somatic and transcriptomic profile of 123 acral melanoma samples from Mexican patients, one of the largest cohorts reported for this type of cancer. In our view, this study helps address several research gaps: 1) The underrepresentation of samples of Latin American ancestry in cancer sample repositories^14^: As it has been shown previously, genetic ancestry and environment influence the somatic profile of tumours, with potential impacts on patient management and treatment^11–13^, 2) the relative lack of studies of acral melanoma, when compared to other types of the disease, as this type of melanoma constitutes the majority of cases in some low- and middle-income countries (LMICs)^3^, and 3) the relative paucity of genomic studies performed and directed from LMICs, such as Mexico.

Most patients in this study had predominantly Amerindian genetic ancestry, which allowed us to perform an analysis of genetic ancestry correlates with somatic mutation profile. We identified a positive correlation between European ancestry and *BRAF* mutation rate (**Figure 1c**). A possible link between European ancestry and *BRAF^V^*^600^*^E^*mutation had been described previously^10^, and this study provides further confirmatory evidence. Other similar correlations have recently been described for other types of cancer, such as a positive relationship between Native American ancestry and *EGFR* mutation rate in lung cancer^13^, and an increased rate of somatic *FBXW7* in African patients compared to European patients^11^. In accordance with this observation, other cohorts of acral melanoma, which studied patients with predominantly European ancestry, have a higher *BRAF* mutation rate than that in this study (*e.g.*, 23% in Australian patients with predominantly European ancestry^7^). These observations should provide the basis for future studies exploring the relationships between ancestry and somatic mutation rate.

We were intrigued to discover that *BRAF*-missense acral melanomas exhibit a more ‘CM-like’ transcriptome than other genetic subtypes of acral melanoma. One possible explanation is that this gene signature is uniquely downstream of a *BRAF* missense mutation. However, in further analyses, we do not see evidence for this explanation (**Figures 3c,d** and **Supplementary Figure 9**). An alternative explanation involves the distinct origins of *BRAF*-missense acral melanomas compared to other acral melanomas. In our previous work^37^, we identified distinct subclasses of human epidermal melanocytes: a common type enriched in limbs (c-type) and a rare type enriched in volar regions (v-type). We observed that most acral melanomas generally retained a transcriptional signature like v-type melanocytes, while a significant subset appeared more akin to c-type melanocytes^37^. The current work indicates that these tumours are more likely to belong to the *BRAF*-activating genetic subtype, suggesting that a subset of volar melanomas might be more accurately classified by cell of origin and/or genetic profile as non-acral CM, rather than bona fide acral melanomas. It is important to clarify that the hypothesis that acral melanomas may arise from different cells of origin is not solely based on this study but is supported by prior work. Our previous research has demonstrated transcriptional diversity among melanocytes in different anatomic locations, including distinct populations of epidermal melanocytes in the palms and soles^37^. Additionally, our zebrafish model studies have shown that AM-associated drivers preferentially (though not exclusively) induce tumours in the limbs (fins), whereas CM-associated *BRAF* mutations preferentially (but not exclusively) lead to tumours in the trunk^38^. Furthermore, we have demonstrated that *BRAF* mutations selectively drive hyperproliferation in only a subset of less-pigmented primary human melanocytes^34^. Therefore, while our additional analyses do not strongly support an oncogene signature as the explanation for the differences in transcriptional scores, thus favouring the cell-of-origin hypothesis, it is possible that in some cases these two phenomena could be intertwined. For example, recent data has shown that some acral melanomas harbouring amplifications of the *CRKL* oncogene depend upon *HOX13* positional identity programs already present in the cell of origin, suggesting that oncogenes and cell-of-origin programs can synergize^38^. Future studies could shed light on this ongoing research area, and could explore the diagnosis of cutaneous melanoma as acral versus non-acral based on molecular signatures rather than solely on anatomic location. The fact that *BRAF*-mutated tumours occur less frequently on patients of non-European ancestry highlights the need to study a diverse set of samples to maximise clinical benefit to all patients. Other observations, such as a tendency for *KIT*-mutated tumours to occur in patients with a higher Amerindian ancestry, are intriguing and will need to be investigated in future studies.

The observation that any mutation in a driver gene (*BRAF*/*NRAS*/*KIT*/*NF1*) leads to worse prognosis is intriguing. Similar observations, namely that mutation in a MAPK pathway gene leads to worse prognosis, have been made in cutaneous melanoma. For example, Long and collaborators reported in 2011 that untreated patients with *BRAF*^V600E^ mutations have worse overall survival than patients with wild-type *BRAF* tumours^39^, an observation that has been replicated in other studies^40–42^, with these observations supporting the use of *BRAF* inhibitors in the clinic^43^. Other mutations in the MAPK pathway also led to shorter overall survival, sometimes in combination with other prognostic factors^44,45^. A previous study also found that mutations in the MAPK pathway predict worse survival in acral melanoma^46^. Regarding *NF1* specifically, previous reports in cutaneous melanoma have suggested that *NF1*-mutated tumours have differing characteristics, sometimes also associated with worse survival and differential response to immune checkpoint inhibition^47,48^. Our results, where we find that patients carrying *NF1* mutations have worse recurrence-free survival than patients with no mutations in this gene, support and expand these previous observations.

Patients with Cluster 1 tumours also showed a better prognosis than other patients, which is not surprising given their associated clinical characteristics (lower Breslow thickness, a tendency for earlier stages at diagnosis, and lower mitotic indexes). However, what is surprising is the gene expression profile characteristic of this cluster. More CAFs and CD4+ T cells were found by deconvolution to be associated to Cluster 1 than other clusters, signatures commonly associated with immunosuppression. A possible explanation is that early-stage tumours are associated with immunosuppressive microenvironments, a balance which, in later tumours, may have been tilted in favour of tumour cell growth. Another potential explanation may involve the recently described roles of CAFs in immunostimulation^49^. Patients with Cluster 2 tumours, with a ‘proliferative/pigmentation’ signature showed the worst survival, with an overexpression of genes associated with proliferation and pigmentation. It has previously been observed in a zebrafish model and in TCGA samples that a pigmentation signature also predicts worse survival^50^, and, in a recent report by Liu and collaborators^25^, AM tumours with a proliferative signature also were associated with worse survival than other tumours. This study both extends and replicates these findings in acral melanoma.

This study has several limitations. Firstly, the nature of our samples (FFPE) may introduce artifacts that affect our ability to identify SNVs and indels accurately. Therefore, to mitigate this risk and increase specificity, we used three different variant calling tools, albeit at the cost of reduced sensitivity. As formalin fixation can generate DNA fragmentation, this may also affect copy number estimation analyses and consequently copy number mutational signature analysis. To mitigate this, we stringently filtered the samples used for this analysis, which affected our statistical power due to a reduced sample size. This study was also done using whole exome data, which limits our ability to call mutations in non-coding regions of the genome.

The characteristics of the analysed cases reflect in part the challenges of setting up such a study. For instance, the fact that the year of diagnosis preceded the date of recruitment by up to 10 years means that somatic mutations associated with higher mortality rates would be under-represented among those recruited, while survival is likely extended over a similarly sized cohort of prospectively recruited cases. To assess the impact of any biases on our interpretation of the impact of mutations, we restricted attention to the 40 tumours diagnosed after mid-2016, *i.e.,* those closest to the time of recruitment. Analysis of survival gave qualitatively and quantitatively similar results to those reported above; in total, 7 of 23 (30.4%) of cases with mutated tumours had died, as opposed to 0 of 17 among cases with QWT tumours. In the analysis of recurrence, again results matched with 70.0% of cases with mutated tumours recurring (16 of 23 cases) compared to 17.7% of QWT tumours (3 of 17, *P* = 0.001). Results for the RNA clusters were similar to the results quoted above for both survival and recurrence.

Overall, we were able to identify novel associations of the germline and somatic profile in AM, genomic-clinical correlates of overall and recurrence-free survival, as well as transcriptional differences in *BRAF-*mutated AMs. This study shows the value of studying diverse populations, allowing us to uncover previously unreported relationships and better understand tumour evolution.

## Methods

A flowchart describing the analyses, steps, and number of samples used in each individual section can be found in **Supplementary Figure 14**.

### Patient recruitment and sample collection

The protocol for sample collection was approved by the Mexican National Cancer Institute’s (Instituto Nacional de Cancerología, INCan, México) Ethics and Research committees (017/041/PBI;CEI/1209/17) and the United Kingdom’s National Health Services (NHS, UK) (18/EE/00076). Patient samples collected for the Utah cohort analysis were derived as previously described^51^.

Recruitment of patients and sample collection took place from 2017 to 2019. Patients attending follow up appointments at INCan that had previously been diagnosed with AM were offered to participate in this study, and upon signing a written consent form, were asked to provide access to a formalin-fixed paraffin-embedded (FFPE) sample of their tumour tissue that had been kept at the INCan tumour bank, as well as a saliva or normal adjacent tissue sample. FFPE samples underwent inspection by a medical pathologist to establish whether sufficient tumour tissue was available for exome sequencing. Saliva samples were collected using the oragene DNA kit (DNAGenotek, # OG-500).

### DNA and RNA extraction

DNA extraction from all saliva samples was performed at the International Laboratory for Human Genome Research from the National Autonomous University of México (LIIGH-UNAM) using the reagent prepITL2P (DNAGenotek, # PT-L2P) and the AllPrep DNA/RNA/miRNA Universal Kit (Qiagen, #80224). DNA and RNA extraction from FFPE samples was performed at the Wellcome Sanger Institute (UK) using the All-prep DNA/RNA FFPE Qiagen kit. Samples with >0.1ng/μl were sequenced through the Sanger Institute’s standard pipeline. Saliva and adjacent tissue samples were used for whole exome sequencing, and only saliva samples were used for genotyping.

### Genotyping

Genotyping was performed using Illumina’s Infinium Multi-Ethnic AMR/AFR-8 v1.0 array at King’s College London and Infinium Global Screening Array v3.0 at University College London. Sufficient germline DNA was available for genotyping for 80 out of 92 samples (86.9%). Ancestry estimation was performed using the ADMIXTURE^52^ v1.3.0 unsupervised analysis together with the superpopulations of the 1000 Genomes dataset. Five superpopulations were identified, corresponding to AFR (Q1), AMR (Q2), SAS (Q3), EAS (Q4), and EUR (Q5) (**Supplementary Table 2**, **Supplementary Figure 1**).

### Exome sequencing and data quality control

FFPE samples, saliva and normal adjacent tissue underwent whole exome sequencing as follows: Exome capture was performed using Agilent SureSelect AllExon v5 probes and paired-end sequencing was performed at the Wellcome Sanger Institute (UK) in Illumina HiSeq4000 machines. Control and tumour samples were sequenced to a mean coverage of 43.72x. Alignment was done using BWA-mem^53^ v0.7.17, using the GRCh38 reference genome. Sequencing quality filters were performed using samtools stats^54^ and fastqc^55^. Sample contamination was estimated using the GATK v4.2.3.0 tool CalculateContamination^56^. Concordance between sample pairs was estimated using the Conpair v0.2 tool^57^. Samples that had <90% similarity with their pair (tumour-normal) or showed a level of contamination above 5% were excluded from the study. After this step, 123 samples remained for further analysis.

### Somatic SNV calling and identification of driver genes and mutations

Somatic variant calling was done using three different tools (CaVEMan^58^ v1.15.2, Mutect2^59^ v4.1.0.0 and Varscan2^60^ v2.3.9), keeping only the variants identified by a minimum of two out of the three tools. For *BRAF*^V600E^ mutations, we kept these variants even if it was only identified by one of the tools as its oncogenicity and relevance in melanoma is well known. When available within the variant calling tool, strand bias filters were applied. A minimum base quality score of 30 on the Phred scale was used. Indel calling was performed using pindel^61,62^ v1.15.2 using indel candidates identified by the structural variant caller manta^63^. When selecting one sample per patient, preference was given to primaries, and metastases or recurrences were chosen only when a primary had not been collected.

Significantly mutated genes were identified using the tool dNdScv^64^ v0.0.1.0 with default parameters using SNVs identified by two of the three tools used for variant calling and indels identified by pindel as input data. Positive selection was considered for genes that had global q-values below 0.1 according to the dNdScv tool recommendations.

Two lymph node metastasis samples (one from a patient that had a *BRAF*^V600E^ mutation and another one with an *NRAS* mutation) and their primaries were annotated as having the same mutation for follow up analysis after manual inspection using IGV^65^.

### Analysis of correlation between driver mutations and clinical covariates and ancestry

Statistical tests were performed to identify potential clinical and ancestry covariates that correlated with driver mutational status. For tumour stage, sex, ulceration status and tumour site, which are discrete variables, association was tested with contingency Chi-squared tests.

For each of the four driver genes, a logistic regression model was fitted to predict the presence or absence of a mutation on the AM samples using the inferred ADMIXTURE^52^ cluster related to the European ancestry component from the 1000 Genomes Project, correcting for age, sex, and tumour mutational burden (TMB, SNVs + indels), as such: Driver gene status ∼ EUR related cluster proportion + age + sex + TMB. Then the log odds related to the EUR cluster were plotted with their respective confidence intervals. The models were constructed using 80 samples out of the 92, which were those with available genotyping information and with all tested covariables available.

### Somatic DNA copy number calling

Sequenza^66^ v3.0.0 and ASCAT^67^ v3.1.2 were used to estimate ploidy and purity values for each sample. These values per sample were compared between the two tools, and samples that had a high discrepancy in their purity estimates (<0.15 vs 1, respectively) were filtered out (**Supplementary Table 28**). Samples with an estimated goodness of fit below 95 were also filtered out. Subsequently, copy number, cellularity and ploidy values estimated by ASCAT were used in follow-up analyses. Whole genome duplication events were assigned as reported by ASCAT. Significantly affected regions by CNAs were identified using GISTIC2^68^ v2.0.23. Amplifications were classified as low-level amplifications when regions had a copy number gain above 0.25 and below 0.9, and as high-level amplifications when regions had a copy number gain above 0.9 according to GISTIC2 values; deletions were classified as low-level deletions when regions had a copy number change between -0.25 and -1.3 and as high-level deletions when regions had a copy number change lower than -1.3. Only peaks with residual q-values < 0.1 were considered as significantly altered. For the analyses of differences in CNA burden by sample group (*e.g.*, mutational status or site of presentation), we used the CNApp tool^69^ to generate copy number alteration scores for global (GCS), focal (FCS) and broad (BCS) CNA burden, calculating segment means (seg.mean) as log2(cn/ploidy) and using default parameters. GCS (Global copy number alteration score) is a number quantifying the copy number aberration level in each sample provided by the CNApp tool^69^. Higher GCS scores indicate a higher burden of copy number aberrations compared with all other samples in the cohort. GCS is the sum of the normalised BCS (Broad copy number alteration score) and FCS (focal copy number alteration score), which are calculated considering broad (chromosome and arm-level) and focal (weighted focal CNAs corrected by the amplitude and length of the segment) aberrations per sample. These values are calculated using as input the number of DNA copies normalised by sample ploidy. A more detailed explanation can be found in the original publication^69^. GCS values were used for comparisons between sample groups. All paired comparisons between groups were evaluated with a Mann-Whitney test.

### Mutational signature analysis

Mutational matrices were generated using SigProfilerMatrixGenerator^70^ v1.2.20. These matrices, with single nucleotide mutations found by at least two of the three variant callers and all insertions and deletions identified by pindel, were used as input for mutational signature extraction using SigProfilerExtractor^71^ v1.1.23 and decomposition to COSMICv3.4 reference mutational signatures^72^. For single base substitutions, the standard SBS-96 mutational context was used, with default parameters and a minimum and maximum number of output signatures being set as 1 and 5, respectively. A total of 116 samples with an SNV count > 0, were used for this analysis. For copy number signature analysis, all 60 samples with available copy number data were used with default parameters, and using the standard CN-48 context from COSMICv3.4.

### RNA sequencing and data quality control

Total RNA library preparation followed by exome capture using Agilent SureSelect AllExon v5 was performed on Illumina HiSeq 4000 machines on 146 samples. Reads were aligned to the GRCh38 reference genome using the splice-aware aligner STAR^73^ v2.5.0. Of these, we focused on the 77 samples that came from different patients, that had matching DNA and were primaries for the score analysis (Methods below). We then applied further quality control filters for the consensus clustering analysis: samples were excluded if total read counts were fewer than 25 million, or if the sum of ambiguous reads and no feature counts was greater than the sum of all gene read pair counts. Forty-four samples remained for downstream analysis. Counts were generated with HTSeq^74^ v0.7.2. Transcripts per million (TPM) normalisation was performed and values were log_2_(TPM+1) transformed.

### Acral vs. non-acral cutaneous tumour score

Patient samples collected for the Utah cohort analysis were derived as previously described^51^. Invasive acral and non-acral cutaneous melanomas were identified and collected as part of the University of Utah IRB umbrella protocol #76927, Project #60, and RNA was extracted and quantified as previously described^51^. A custom NanoString nCounter XT CodeSet (NanoString Technologies) was designed to include genes differentially expressed between glabrous and non-glabrous melanocytes^37,38^. Sample hybridization and processing were performed in the Molecular Diagnostics core facility at Huntsman Cancer Institute. Data were collected using the nCounter Digital Analyzer. Raw NanoString counts were normalised using the nSolver Analysis Software (NanoString Technologies). Normalisation was carried out using the geometric mean of housekeeping genes included in the panel (**Supplementary Table 16**). Background thresholding was performed using a threshold count value of 20. Fold change estimation was calculated by partitioning by acral vs. cutaneous melanoma. The log2 normalised gene expression data were subjected to Principal Component Analysis (PCA) using the PCA function in Prism version 10.2.1 (GraphPad Software). PCA was performed to identify the main sources of variability in the data and to distinguish between acral and cutaneous samples.

To determine the top differentially expressed genes contributing to the variance between acral melanomas and cutaneous melanomas, the loadings of the second principal component (PC2) were examined. Genes with the highest positive and negative loadings on PC2 were selected as the top 10 and bottom 10 genes, respectively. Log2 expression values of these genes were used to generate a multiplicative score, producing the ratio of acral to cutaneous melanocyte genes. Statistical analyses were performed using Prism version 10.2.1 (GraphPad Software). Differences in acral to cutaneous ratios were assessed using the Mann-Whitney U test.

The acral:cutaneous (A:C) ratio was calculated for each of the 77 primary acral tumours using the method described above after batch correction (limma v3.64.1) on normalised and transformed expression data processed by the R package DESeq2 v1.48.1. Differences in the A:C gene expression ratio scores between *BRAF* activating mutation-positive and *BRAF-*wildtype acral melanoma samples were assessed using a Mann-Whitney U test. The same normalisation, scoring method and statistical testing was applied to the 63 transcriptomes from acral melanoma tumours considering *BRAF*-activating (n=10) and wild-type (n=53) in Newell *et al* (2020)^7^. All available samples in this cohort were used, as only one primary had a *BRAF* mutation. Only samples with *BRAF*-activating mutations (V600E and L597R for the Mexican acral melanoma set) were included in the *BRAF* group.

To determine whether the CM classifier genes are induced by *BRAF*^V600E^ signalling in melanocytes, we analysed RNA-seq data from McNeal *et al*^34^, which consisted on bulk-RNA sequencing of primary human melanocytes transduced with *BRAF*^V600E^ and cultured under two conditions: PMA (phorbol 12-myristate 13-acetate) and ET1 (endothelin-1). We extracted normalised expression values for CM classifier genes across four conditions: PMA, PMA + *BRAF*^V600E^, ET1, and ET1 + *BRAF*^V600E^. Normalised expression levels were compared using the Mann-Whitney U test in Prism version 10.2.1 (GraphPad Software).

We evaluated the A:C classifier in clinical melanoma samples using RNA-seq data from The Cancer Genome Atlas (TCGA) Skin Cutaneous Melanoma Firehose Legacy cohort. Normalised gene expression data were downloaded from cBioPortal. Samples were classified as *BRAF*-activating or *BRAF*-wildtype in the same way as for Figure 3C. We calculated the product of the expression of CM classifier genes for each category. Differences were assessed using the Mann-Whitney U test.

We used an interactive Shiny application, What Is My Melanocytic Signature (WIMMS; https://wimms.tanlab.org) to compare transcriptional programs associated with distinct melanocytic cell states. WIMMS classifies melanocytic gene expression profiles by aggregating previously published gene expression signatures and clusters them into seven principal cell state categories. We input our gene signature into WIMMS to assess correlation with these reference states. The resulting dendrogram represents the relationship between our classifier-derived CM (cutaneous melanoma) genes and known signatures.

### Consensus clustering and deconvolution based on gene expression

To identify molecular subgroups based on transcriptome data, we performed consensus clustering using the Cola R v2.10.1 package^75^. Standard preprocessing of the input matrix was performed, including removal of rows in which >25% of the samples had NA values, imputation of missing values, replacement of values higher than the 95th percentile or less than 5th by corresponding percentiles, removal of rows with zero variance, and removal of rows with variance less than the 5th percentile of all row variances. Subsequently, standard statistical metrics were used to assess the number of clusters and the stability of the partitions, including 1-PAC score, concordance and jaccard index, and visual inspection of the consensus matrix through heatmaps visualisations. Afterwards, signature analysis and functional enrichment on the identified clusters were performed.

The EPIC algorithm^76^ v1.1.7 was used in the R programming environment to perform deconvolution in order to infer immune and stromal cell fractions within AM tumours. We used the TRef signature method with default parameters, which includes gene expression reference profiles from tumour-infiltrating cells. The algorithm generated an absolute score that could be interpreted as a cell fraction.

### Survival analyses

Consenting and recruitment of patients started in 2017 and ended in 2019. Because of the challenges of recruiting significant numbers of participants with AM, patients diagnosed in earlier years who were still attending follow-up clinics for primary or recurrent disease were recruited. To ensue data consistency, only participants with a primary available for analysis were the subject of focus on analyses of recurrence and/or death (n=85 patients). In total, 73 participants were recruited whose primary and ancestry data were available for analysis on driver mutations. A total of 44 patients with primary and RNA cluster data available were used for analysis on clusters regardless of their ancestry data availability.

To compare the effect of distinct driver mutations and the RNA clusters, we examined 2 measures of disease severity (i) the recurrence of the primary and (ii) overall survival. For recurrence, we examined time to recurrence using a life-table approach from date of primary diagnosis onwards as a descriptor, and based analyses of differences between mutations (and clusters) on logistical regression adjusting for date of diagnosis, age at diagnosis, sex, stage at diagnosis (advanced/early), ancestry (only for mutations) and time from diagnosis through either death or last known to be alive. Primary tumours were either classified as QWT or mutated for known drivers; tumours with multiple driver mutations were classified as “multihit”. We also conducted analyses based on a binary exposure of “QWT” or “Mutated” tumour based on the existence of one or more mutations in a known driver gene.

For survival analysis, we also included a life-table approach, again as a descriptor, based on time from diagnosis through to date of death or date last known to be alive. Statistical assessment of the effect of each mutation and/or cluster were based on Cox proportional hazard analysis with followup starting from date of recruitment through to date of death (or last date alive) adjusting for date of diagnosis, age at diagnosis, sex and ancestry (as a sensitivity analysis). Analysis of combined mutations were also conducted as for the recurrence analysis.

## Data Availability

Sequencing data are available at the European Genome-Phenome Archive (EGA). DNA sequencing data are available under ENA accession number EGAS00001003740 and RNA sequencing data under ENA accession number EGAS00001003758. Code is available at https://github.com/CGBio-Lab/Mex-acral-exomes-transcriptomes.

---

**Supplementary Figure 1.**
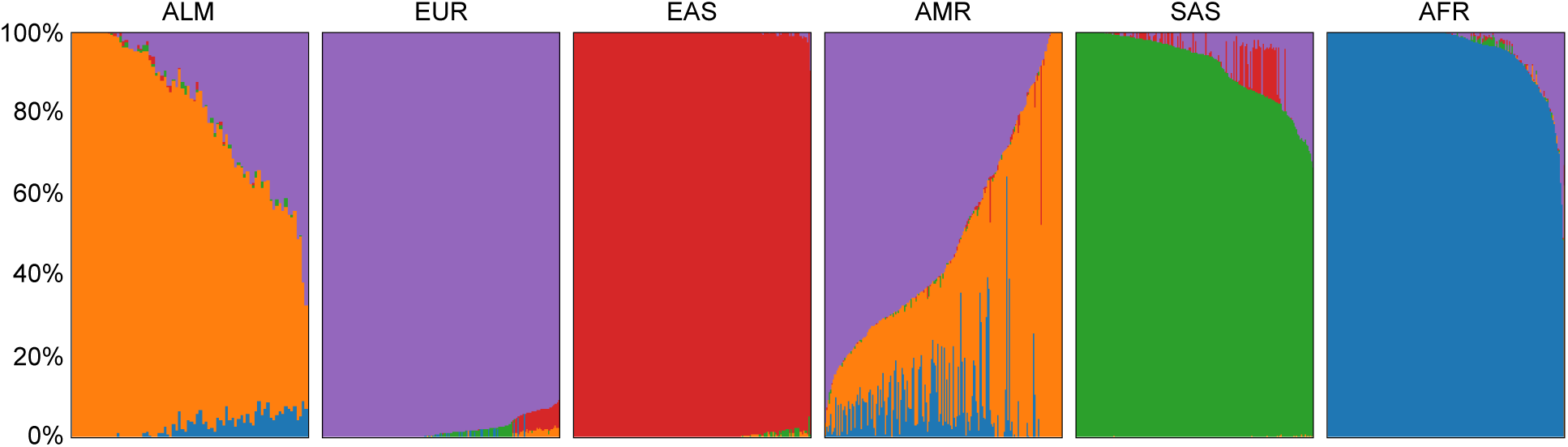
Estimation of ancestry proportions per sample together with the superpopulations of the 1000 Genomes dataset. The leftmost panel corresponds to the samples genotyped in this study (n=80). The following panels correspond to the superpopulations in the 1000 Genomes Project. Five superpopulations are plotted, corresponding to African (AFR, blue), American (AMR, orange), South Asian (SAS, green), East Asian (EAS, red), and European (EUR, purple).

**Supplementary Figure 2.**
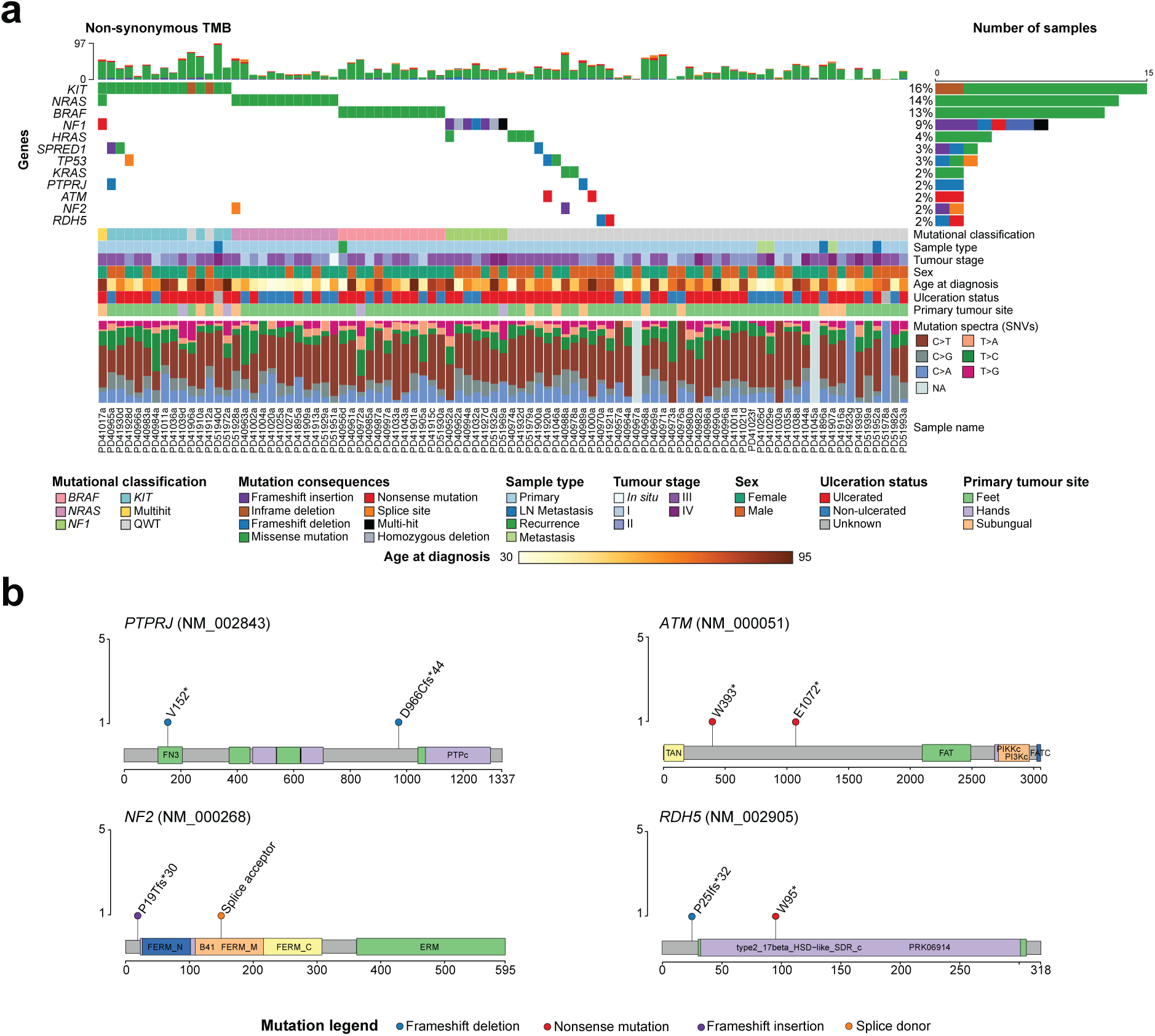
Somatic landscape of all acral melanoma samples. a) Oncoplot depicting the seven most mutated genes according to dNdScv and five selected genes based on mutational frequency and biological function. Mutational classification, sample type, tumour stage, sex, age at diagnosis, ulceration status, tumour site and mutational spectra are shown by sample. b) Mutations found in *PTPRJ*, *ATM, NF2* and *RDH5*, for which all mutations are deleterious and are found altered each in two samples.

**Supplementary Figure 3.**
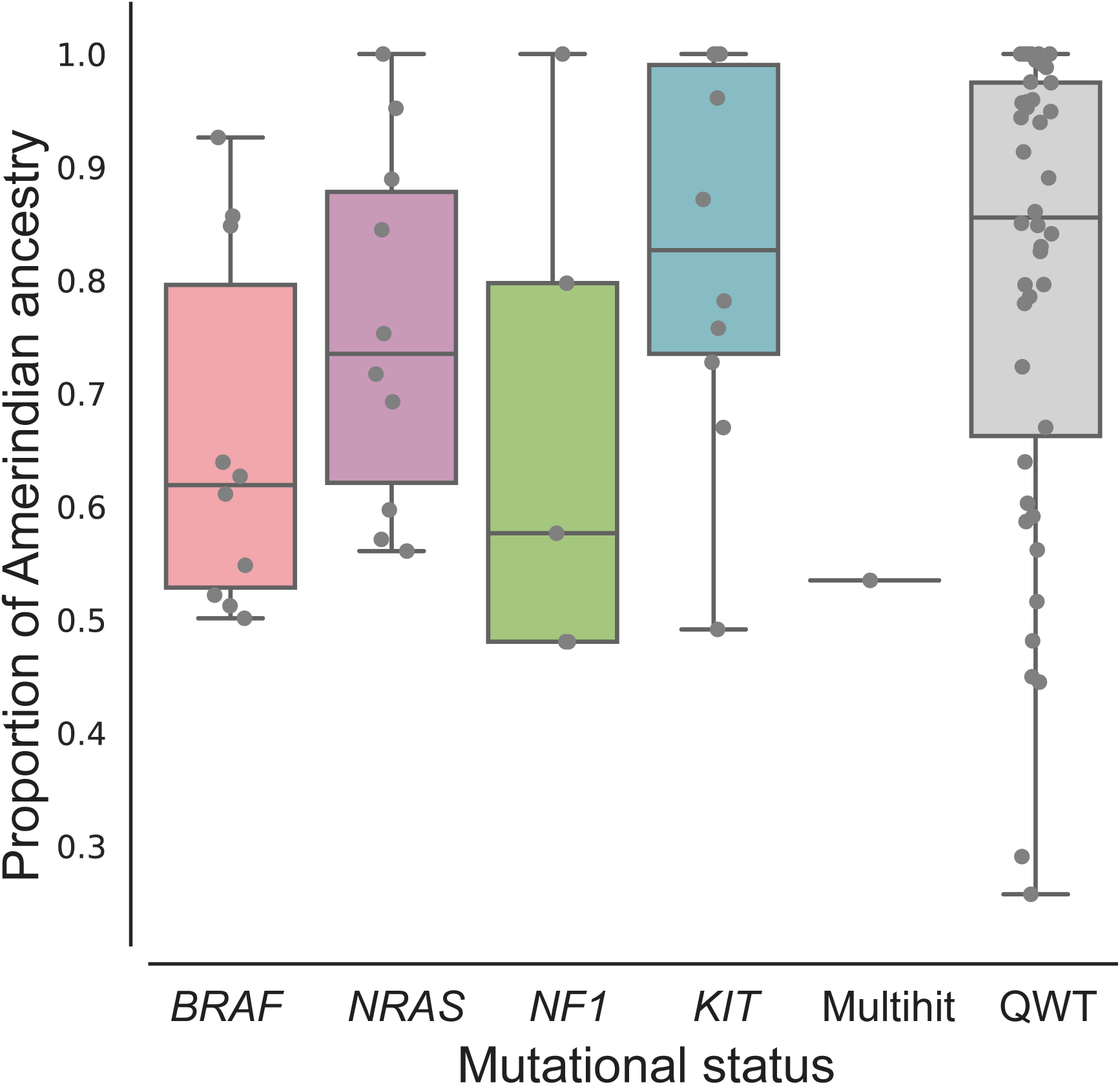
Boxplot of the proportion of Amerindian ancestry among patients classified by genomic subtype. Each dot corresponds to a sample. The central line within each box represents the median value, the box boundaries represent the interquartile range (IQR), and the whiskers extend to the lowest or highest data point still within 1.5xIQR.

**Supplementary Figure 4.**
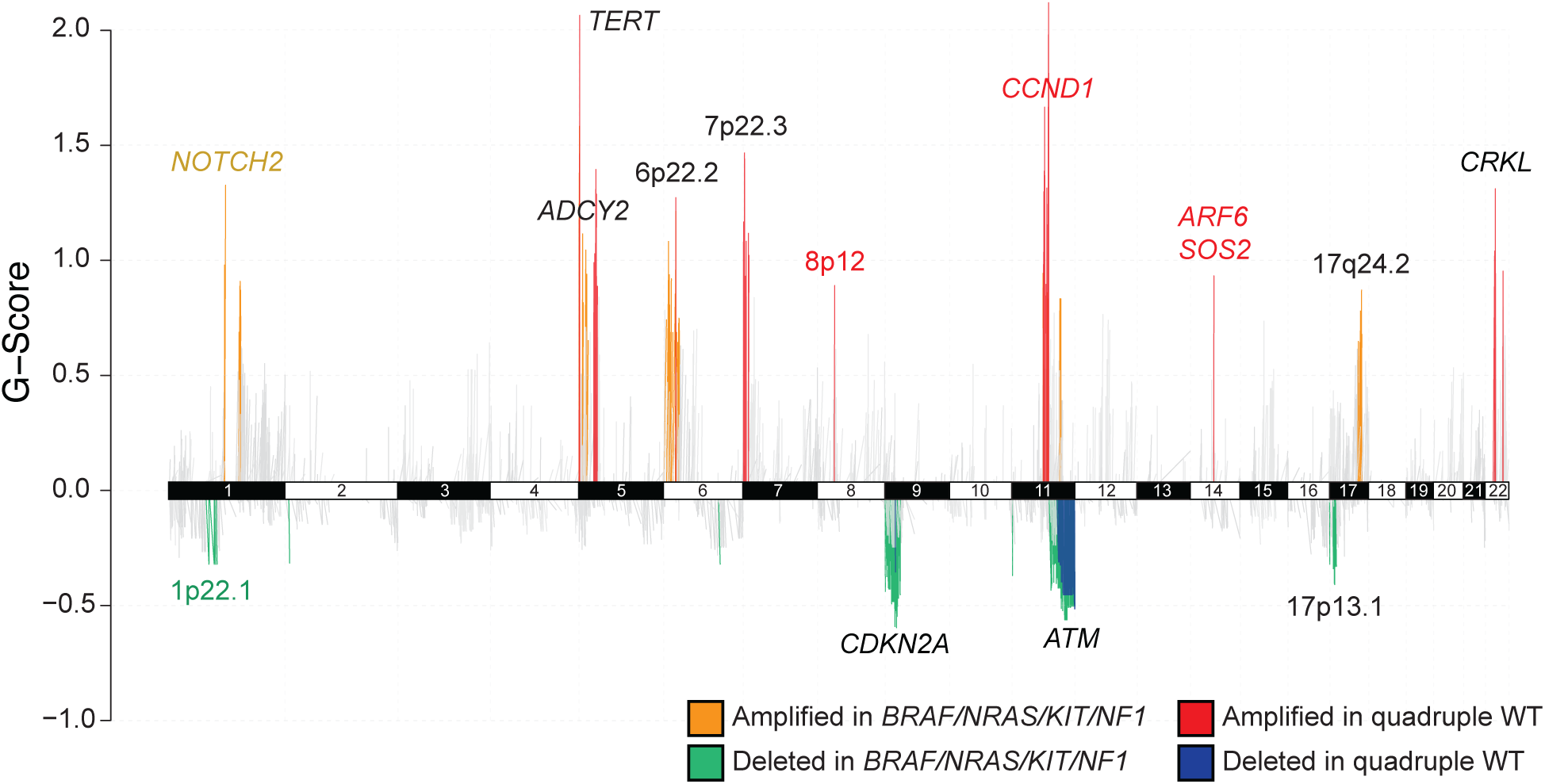
Copy number profile of *BRAF/NRAS/KIT/NF1*-mutated tumours vs. quadruple WT. All depicted regions have been identified by GISTIC2 analysis per group of samples. Statistically significant differences are marked in colour, yellow = amplified in mutated tumours, red = amplified in QWT, green = deleted in mutated tumours, blue = deleted in QWT tumours. Differences are determined first by assessing the global GISTIC2 output and determining differences between groups by one-sided Fisher’s exact test (*P* < 0.05). If a region is not found in the global GISTIC2 output, but it is found only in the analysis per group, we have indicated it as statistically different. Number of mutated tumours = 22, number of QWT tumours = 25.

**Supplementary Figure 5.**
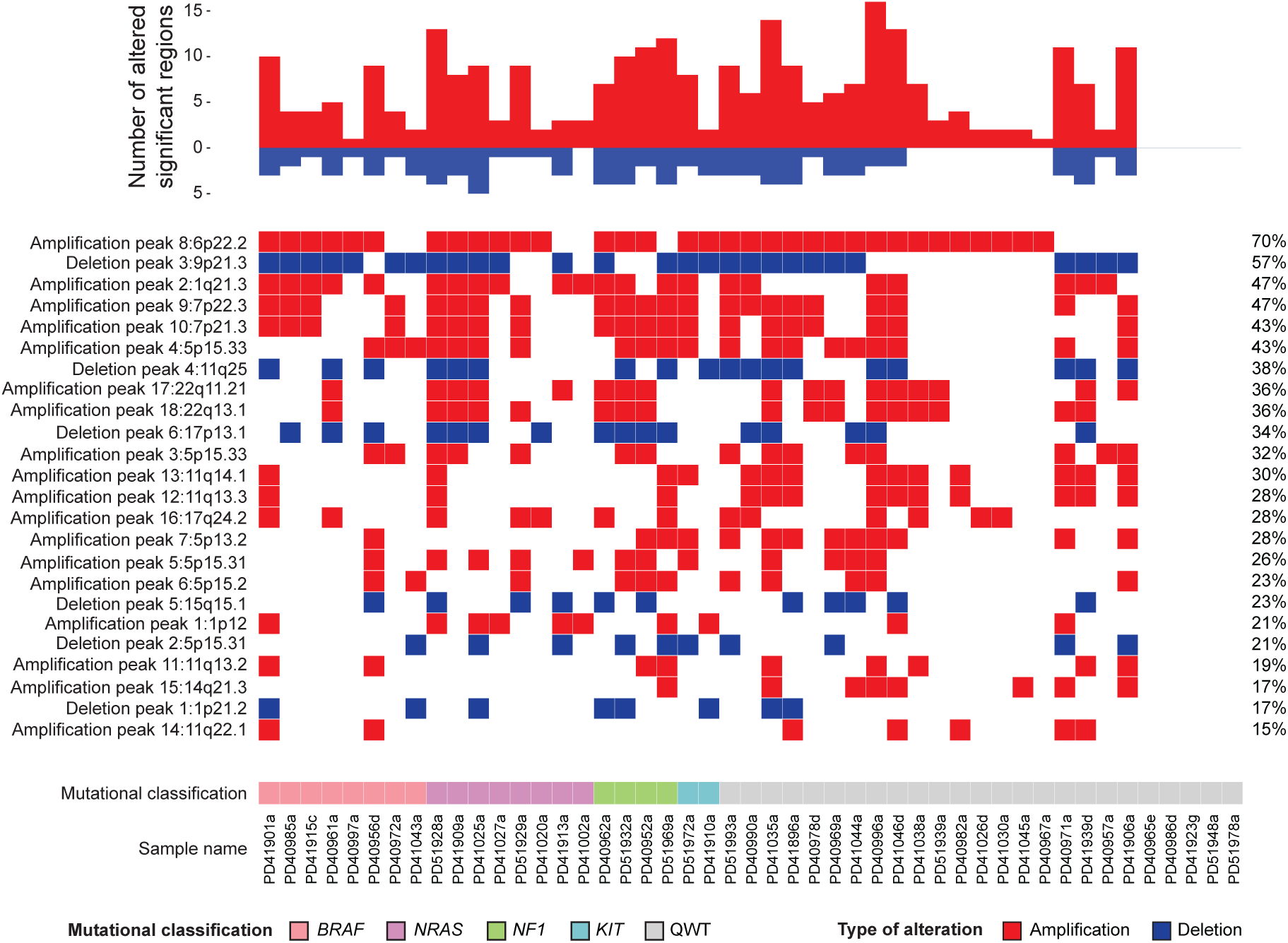
Significantly altered regions per sample. Top panel. Number of significant regions altered by sample. Bottom panel. Binary heatmap showing the significantly altered regions identified by GISTIC2 per sample. One sample per patient is shown. Heatmap is ordered on the X axis by mutational classification and on the Y axis by frequency of alterations per region.

**Supplementary Figure 6.**
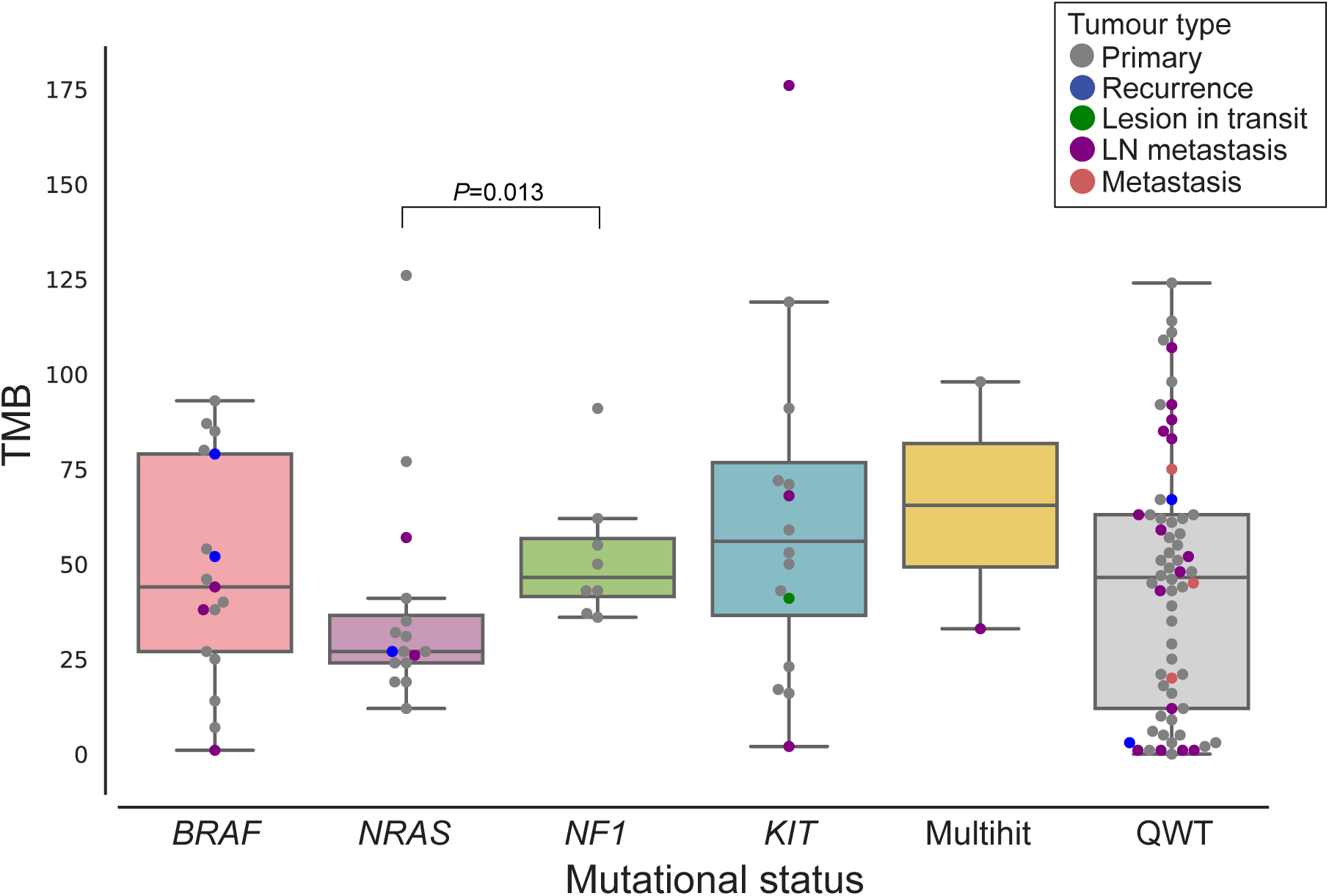
Boxplot of TMB for all samples classified by genomic subtype. Each dot corresponds to a sample, and colours represent tumour type. The central line within each box represents the median value, the box boundaries represent the interquartile range (IQR), and the whiskers extend to the lowest or highest data point still within 1.5xIQR. *P*-values shown are from Wilcoxon Mann-Whitney tests.

**Supplementary Figure 7.**
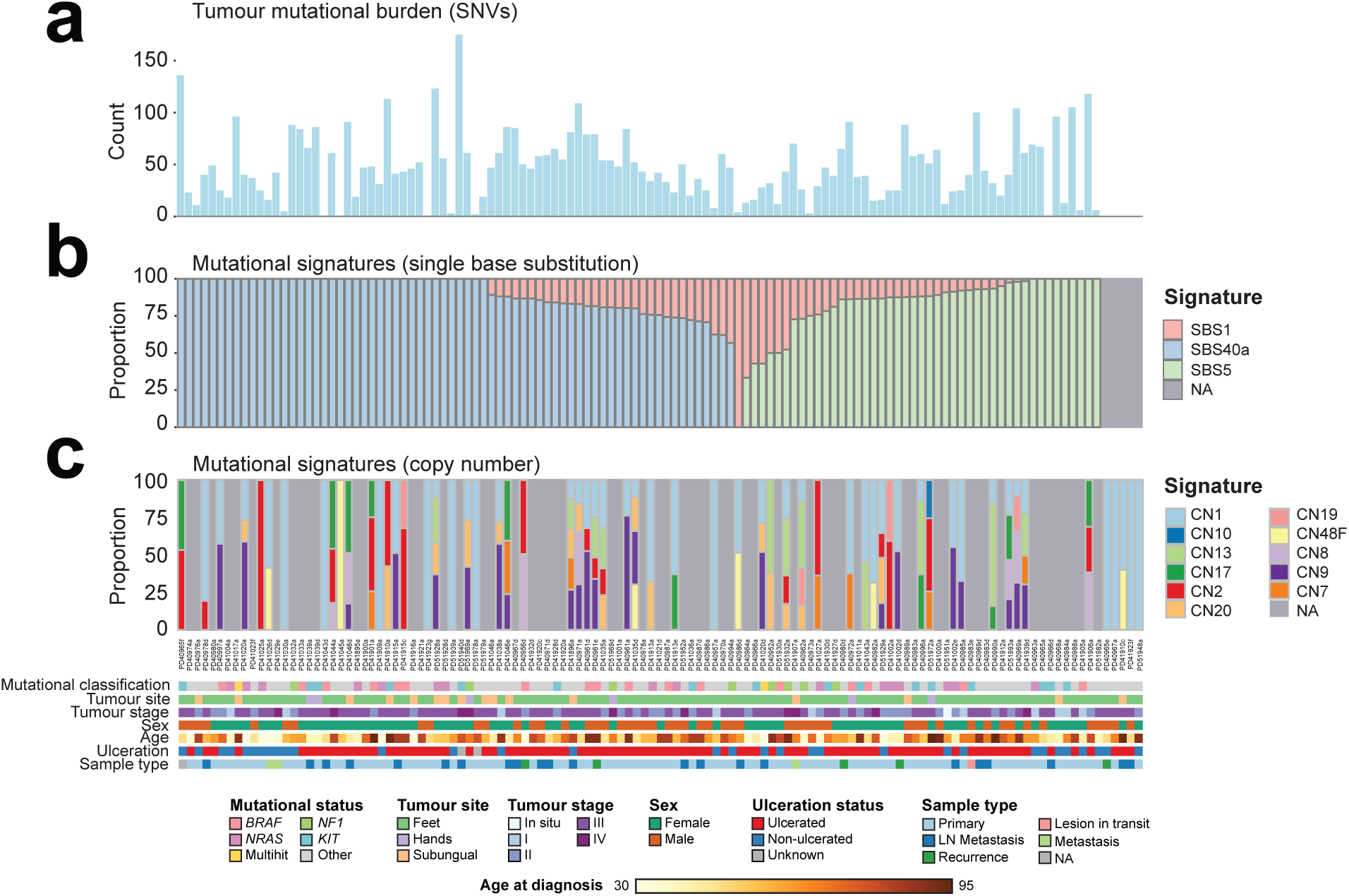
Mutational signatures found in acral melanoma samples from Mexican patients. a) The SNV component of tumour mutational burden per sample. b-c) Proportions of mutational signatures per sample are shown in stacked bars for single base substitutions (b), and copy-number aberrations (c). In b) and c), samples with a light gray background did not have data available. Genomic subtypes and clinical characteristics are plotted at the bottom.

**Supplementary Figure 8.**
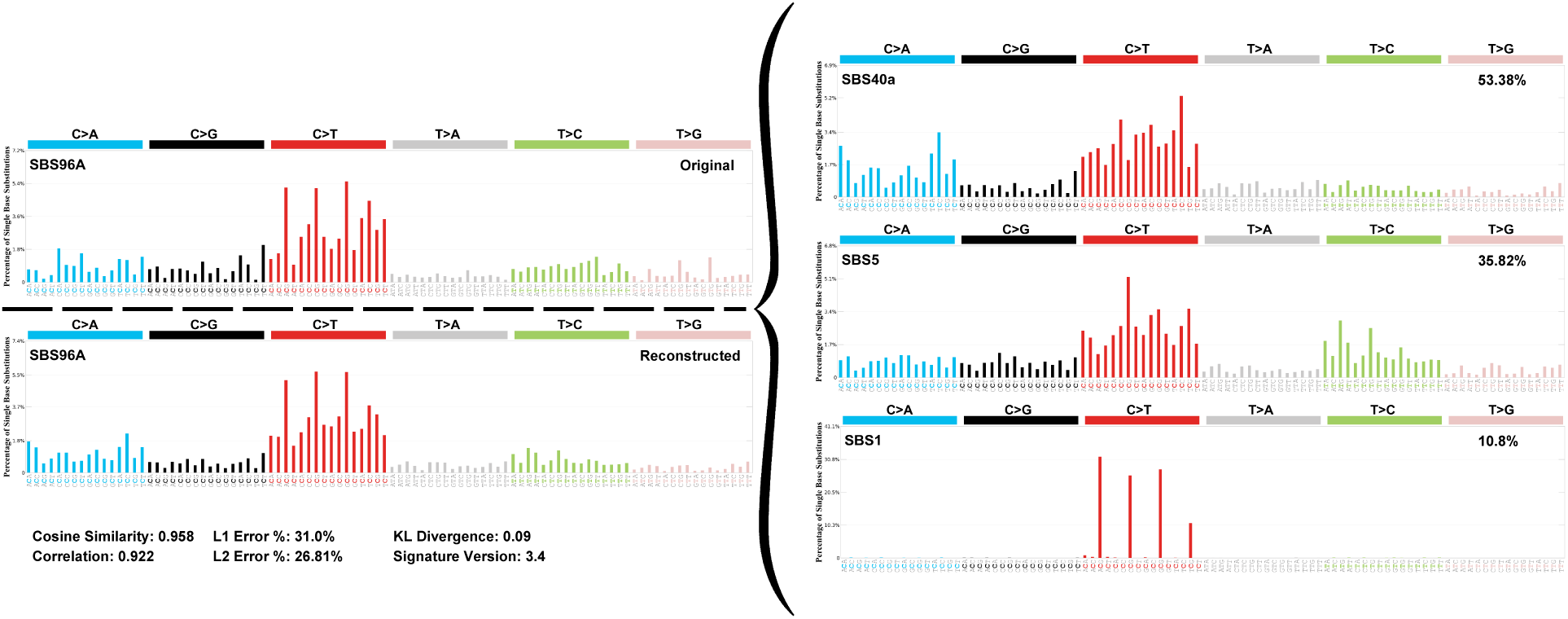
Decomposition plot of single-base substitution mutational signatures for all acral melanoma samples. Samples that had mutation data (116) were included in this analysis.

**Supplementary Figure 9.**
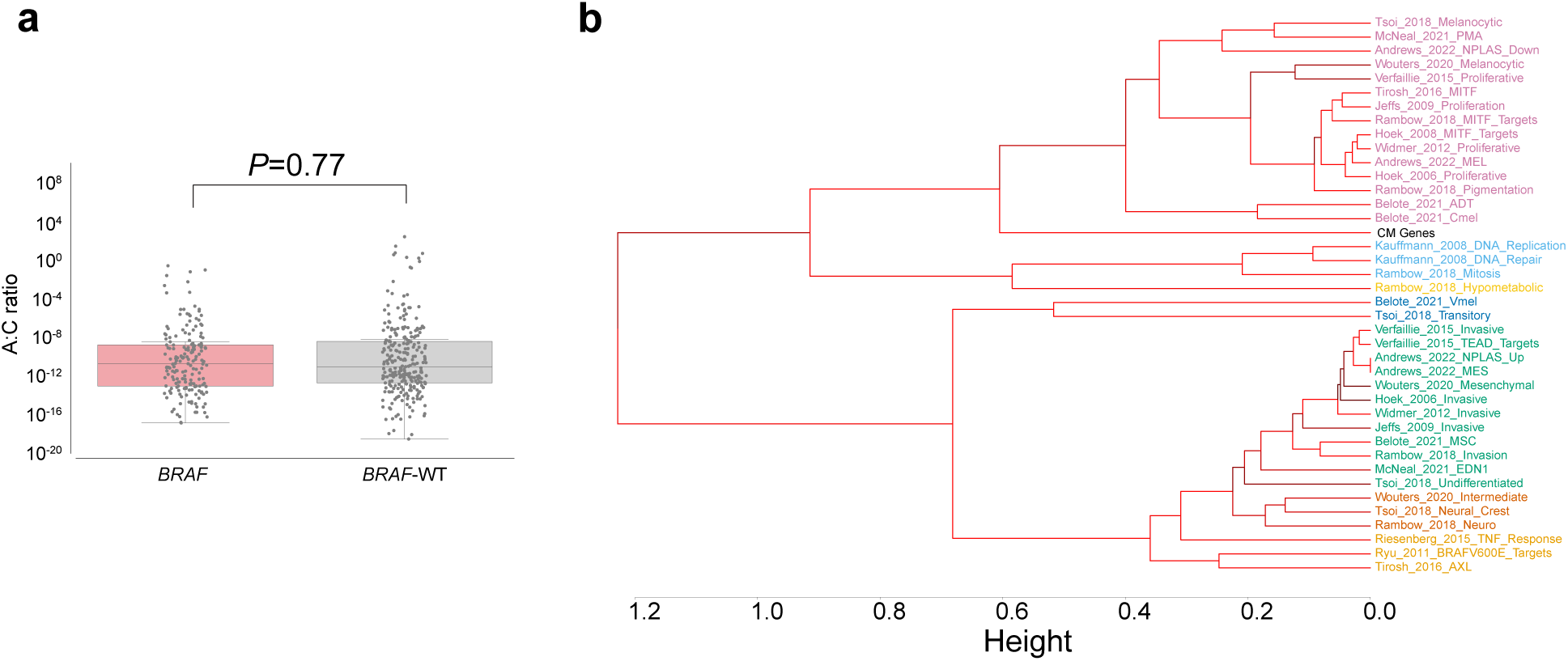
Analyses testing the association of the transcriptional signature found in acral *BRAF*-mutated tumours with downstream *BRAF* signalling. A) Scatter plot comparison of Acral:Cutaneous gene expression ratio in cutaneous melanoma samples from The Cancer Genome Atlas (TCGA) stratified by activating mutation status. Samples were compared in two groups: non-*BRAF* activated tumours and *BRAF*-activating tumours. Statistical significance was assessed using individual Mann-Whitney U test. B) Hierarchical clustering dendrogram generated using the WIMMS platform to compare the cutaneous melanoma classifier genes to other published molecular signatures, including a signature of genes activated by mutant *BRAF*^V600E^ in melanoma cells (Ryu_2011_BRAFV600E_Targets)^36^.

**Supplementary Figure 10.**
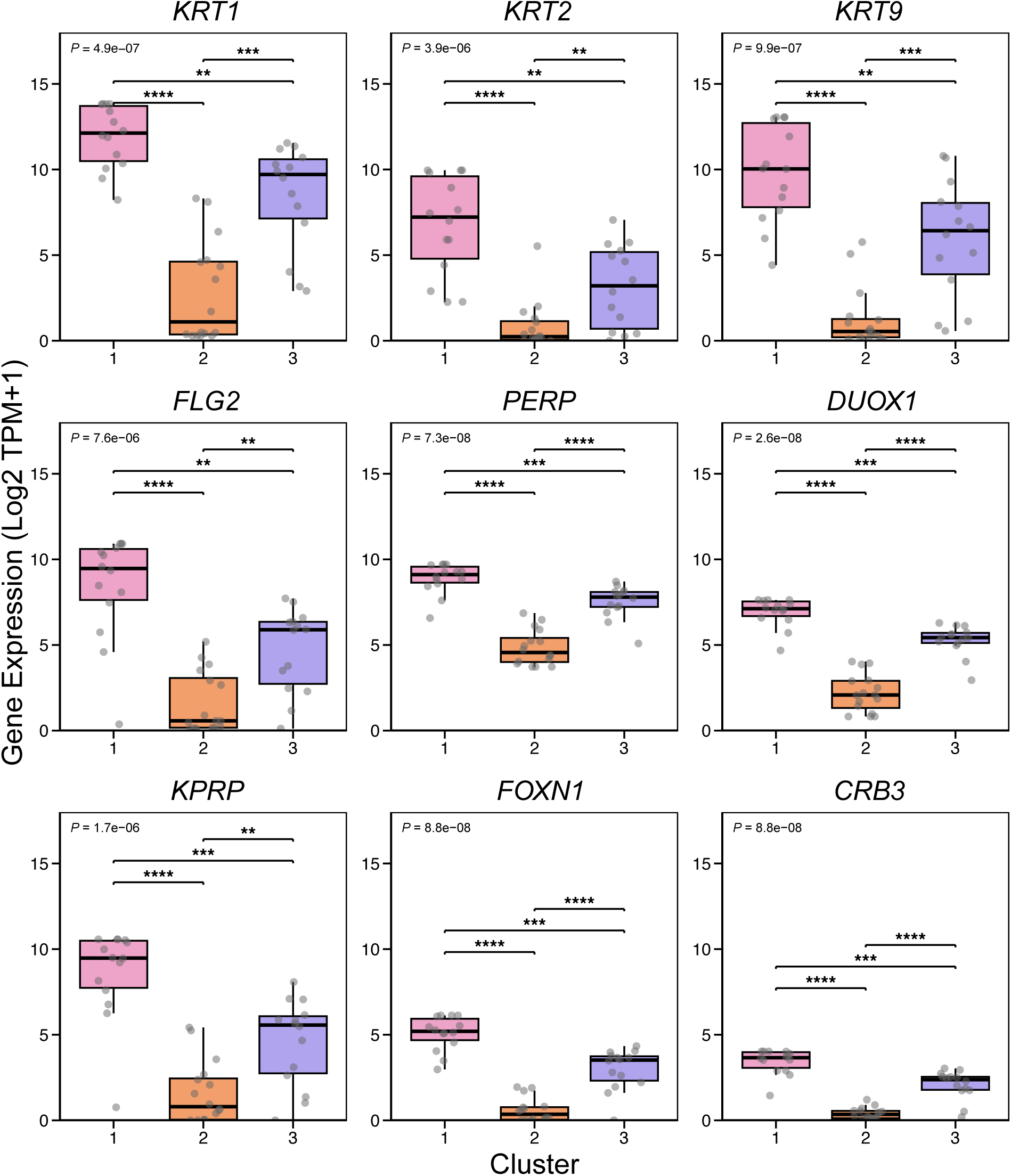
Gene expression for a selection of genes found associated to Cluster 1. *P*-values were estimated with Kruskal-Wallis tests.

**Supplementary Figure 11.**
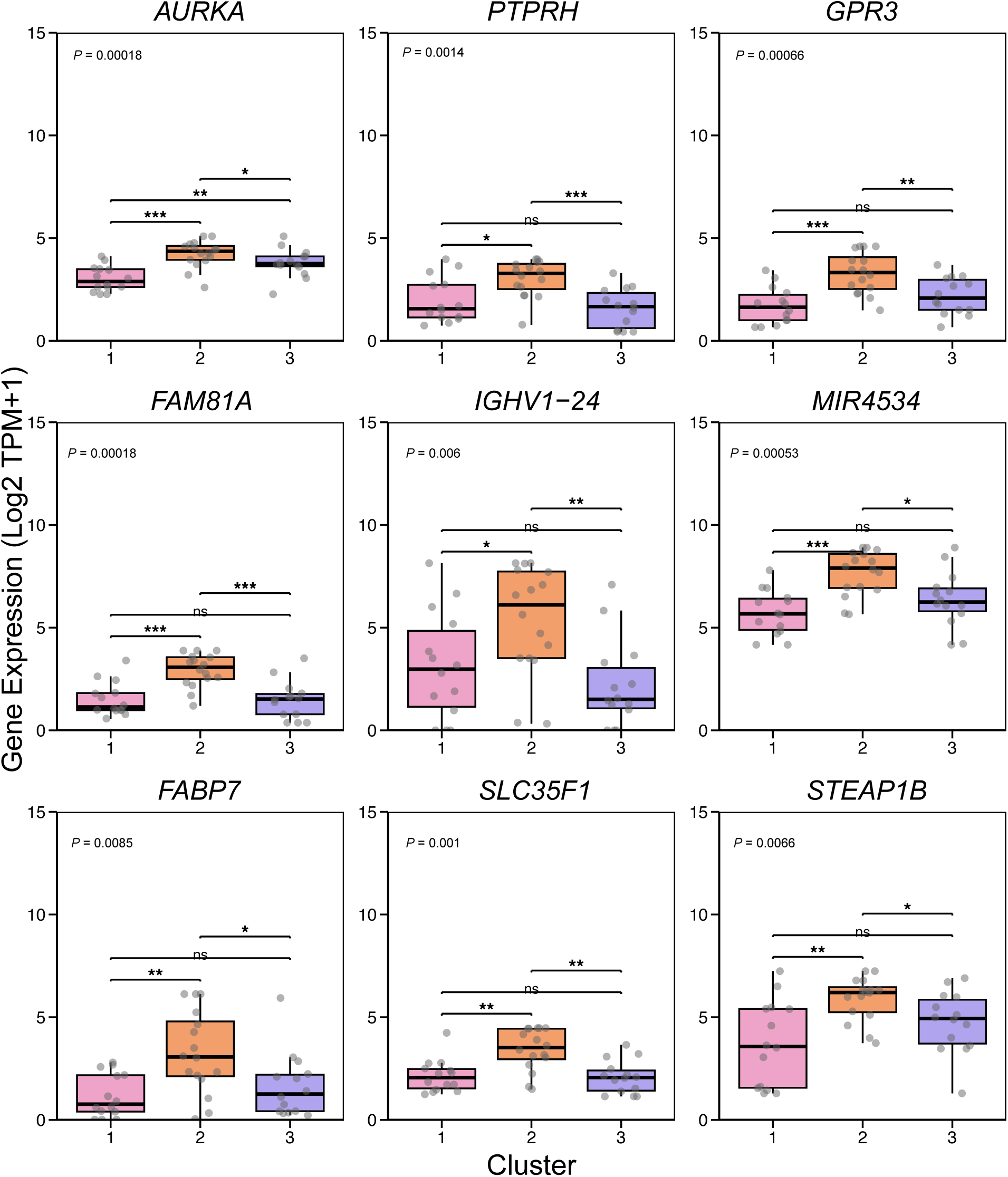
Gene expression for a selection of genes found associated to Cluster 2. *P*-values were estimated with Kruskal-Wallis tests.

**Supplementary Figure 12.**
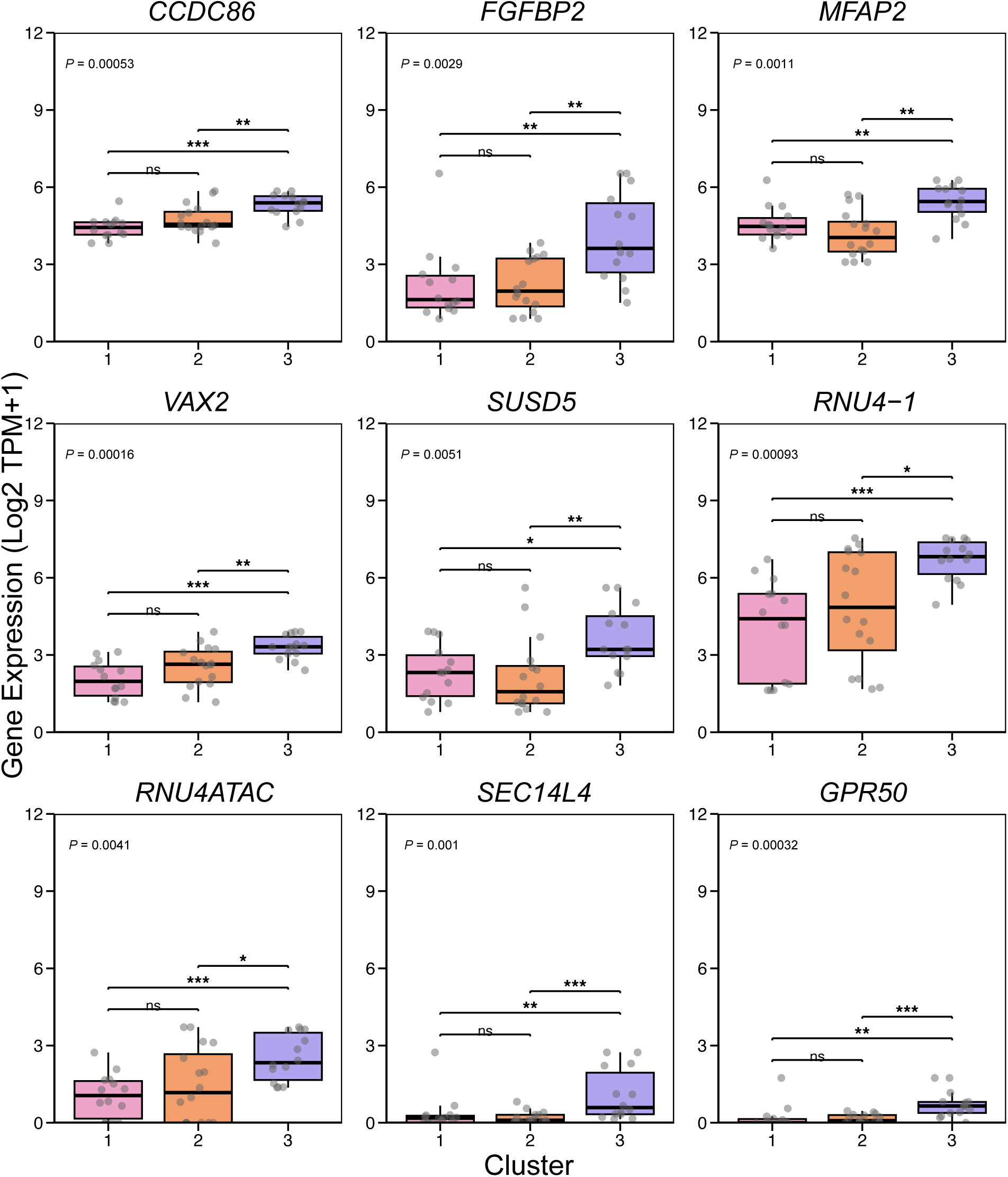
Gene expression for a selection of genes found associated to Cluster 3. *P*-values were estimated with Kruskal-Wallis tests.

**Supplementary Figure 13.**
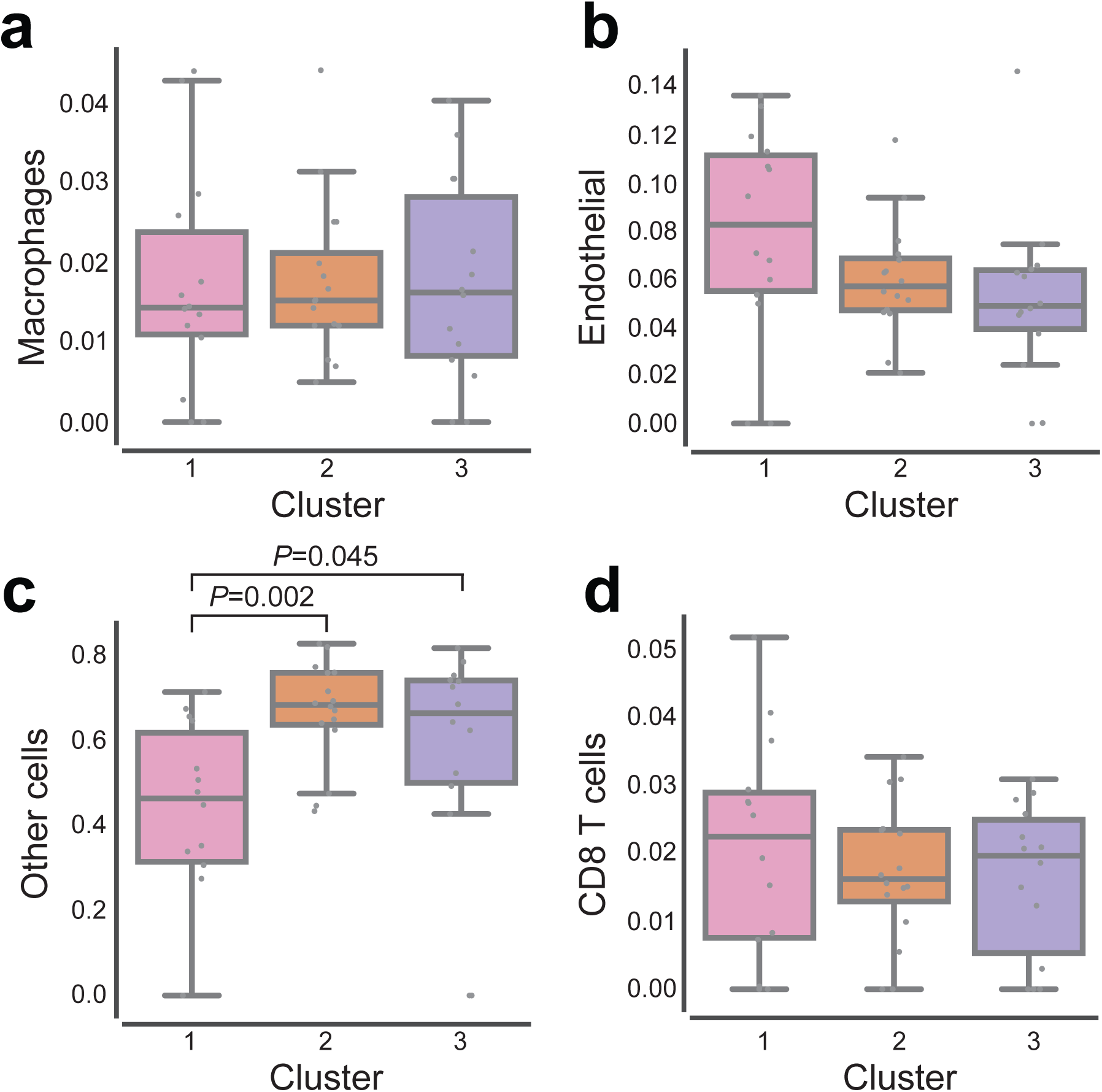
Deconvolution results for macrophages, endothelial cells, CD8 T cells and other cells. A) Box plot of macrophage proportion (Y axis), as calculated by deconvolution, per sample classified by transcriptional cluster. B) Box plot of endothelial cell proportion (Y axis), as calculated by deconvolution, per sample classified by transcriptional cluster. C) Box plot of other cells (non-immune), as calculated by deconvolution, per sample classified by transcriptional cluster. D) Box plot of CD8 T cell proportion (Y axis), as calculated by deconvolution, per sample classified by transcriptional cluster. The central line within each box represents the median value, the box boundaries represent the interquartile range (IQR), and the whiskers extend to the lowest or highest data point still within 1.5xIQR. Individual data points are plotted as dots. Wilcoxon-Mann-Whitney paired tests were performed.

**Supplementary Figure 14.**
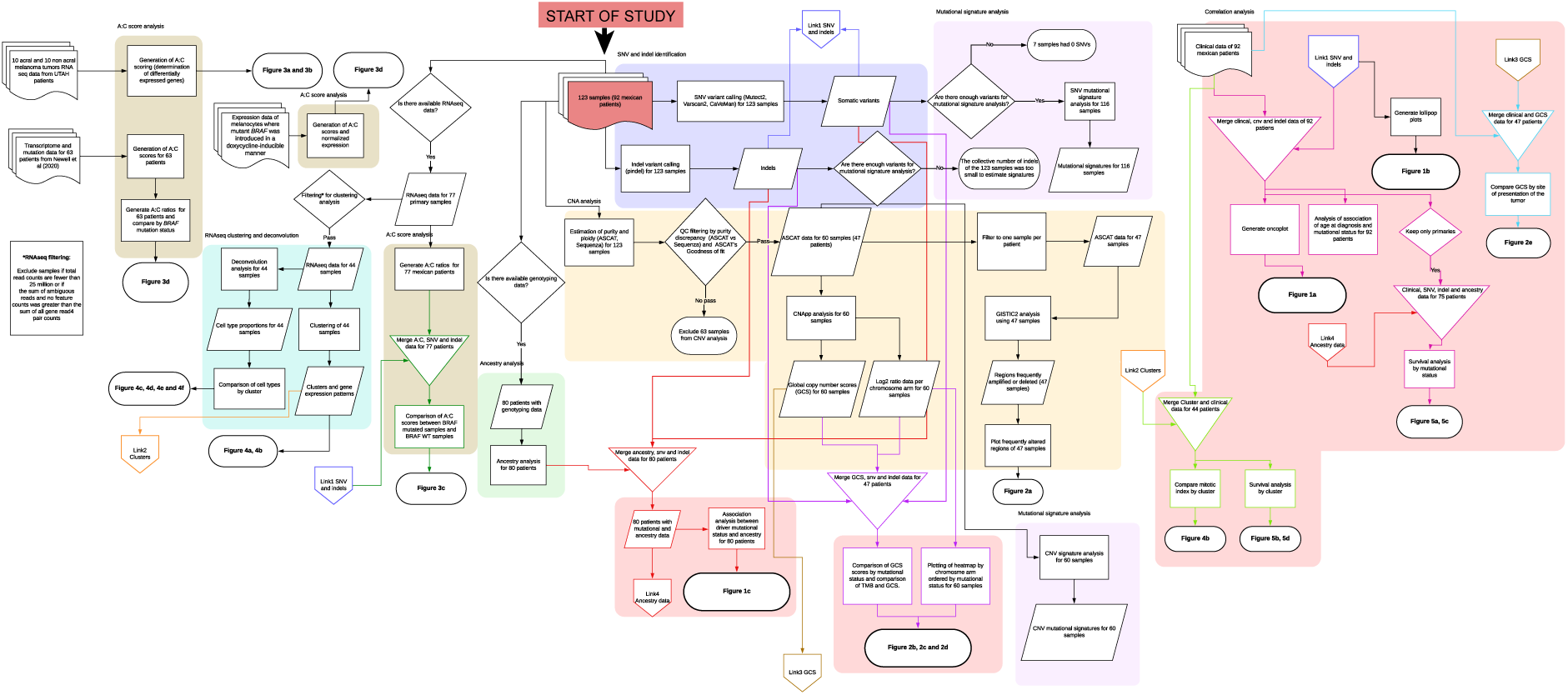
Flowchart describing all analyses and steps in this work. Different colours represent different sections of the study, the resulting main figure from each analysis is also indicated in the text.

## Acknowledgments

We are deeply grateful to patients and their families for agreeing to form part of this study and providing access to their samples. We are also thankful to members of the CGBio lab team at LIIGH-UNAM for valuable discussions regarding the findings in this article. The authors wish to thank Luis A. Aguilar, Alejandro de León and Alejandro Avalos from the Laboratorio Nacional de Visualización Científica Avanzada and Jair S. García Sotelo, Abigayl Hernández, Eglee Lomelín, Iliana Martínez, Rebeca Muciño, María A. Ávila, Christian Molina-Aguilar, Alejandra Castillo and Carina Uribe Díaz from the International Laboratory for Human Genome Research, National Autonomous University of Mexico. We are grateful to the International Cancer Genome Consortium Data Access Committee for granting access to ICGC controlled data. We are also thankful to Kim Wong and Daniel Desposorio for useful discussions. Work included in this paper has been funded by Wellcome Trust (204562/Z/16/Z and 227228/Z/23/Z to C.D.R.-E.), the Melanoma Research Alliance (Pilot Award #825924, to C.D.R.-E.), the Mexican National Council of Humanities, Science and Technology (CONAHCYT, FOSISS A3-S-31603, to C.D.R.-E.), Programa de Apoyo a Proyectos de Investigación e Innovación Tecnológica (PAPIIT UNAM) (IN209422 to C.D.R.-E.), and the Wellcome Sanger Institute through an International Fellowship. This project was also supported by the MRC Dermatlas project; MR/V000292/1. A.J., D.C.D., and R.L.J.-T. are supported by the Department of Dermatology and the Huntsman Cancer Foundation. This work was funded in part by the Melanoma Research Alliance Dermatology Fellows award to D.C.D., the Harry J Lloyd Charitable Trust Melanoma Research Grant to R.L.J.-T., a National Cancer Institute R01 (R01CA229896) to R.L.J.-T., and pilot funds from the Huntsman Cancer Institute Melanoma Center. We utilised the Shared Resources for Research Informatics and High-Throughput Genomics and Bioinformatics Analysis, each supported by the National Cancer Institute of the National Institutes of Health under Award Number P30CA042014. M.D.-G. and P.G.-G. were awarded fellowships within the “Generación D” initiative, Red.es, Ministerio para la Transformación Digital y de la Función Pública, for talent attraction (C005/24-ED CV1), funded by the European Union NextGenerationEU funds, through PRTR. P.B. is a PhD student from Programa de Doctorado en Ciencias Biológicas, Universidad Nacional Autónoma de México (UNAM), and was supported by Consejo Nacional de Humanidades, Ciencia y Tecnología (CONAHCyT, now known as SECIHTI) (holder no. 562546, scholarship no.762536). This paper is part of P.B.’s requirements for obtaining a Doctoral degree at the Posgrado en Ciencias Biológicas, UNAM.

## Author Contributions

P.B.-L., M.E.V.-C., D.C.-I., E.F.R., J.B., P.A.J., I.S.-W., J.R.C.W.-R., K.L.C.-R., A.J., D.C.D., J.I.R.-G., O.I.G.-S. and M.C.V.H. performed bioinformatic and statistical analyses. C.M.-A., F.G.A.-G., M.C.-V., R.O.-L. and L. v.d.W. did sample cataloguing and nucleic acid extraction. E.T.D. provided computational resources and advice on statistical analyses. A.A.-C., D.Y.G.-O., H.M.-S., R.R.-M., H.V.C., L.A.T.-P. and D.H.-U. assessed patients and provided access to biological samples. A.H.-M. provided facilities for sample processing and supervised that part of the work. M.J.A., I.F. and M.T. performed sample histopathology. P.G.G., M.D.-G. and L.B.A. supervised the mutational signatures analysis. Y.S.-P. provided access to patient clinical information and supervised that part of the work. G.K.I., R.L.B. and R.M.W. provided data and information that crucially helped the interpretation of the results in this manuscript. D.T.B. performed survival statistical analyses. P.A.P., R.L.J.-T., D.J.A. and C.D.R.-E. jointly supervised this work. C.D.R.-E. wrote the manuscript with assistance from P.B., P.A.P., R.L.J.-T. and D.J.A.

